# What is the effectiveness and cost-effectiveness of interventions in reducing the harms for children and young people who have been exposed to domestic violence or abuse: a rapid review

**DOI:** 10.1101/2023.05.10.23289781

**Authors:** Llinos Haf Spencer, Annie Hendry, Abraham Makanjuola, Kalpa Pisavadia, Jacob Davies, Mohammed Albustami, Bethany F Anthony, Clare Wilkinson, Deborah Fitzsimmons, Dyfrig Hughes, Rhiannon Tudor Edwards, Ruth Lewis, Alison Cooper, Adrian Edwards

## Abstract

Children and young people witnessing domestic violence and abuse (DVA) can be affected negatively in terms of their psychological, emotional, and social development.

Adverse events in childhood are known to be harmful to a young persons development and influence their life course, and therefore is a significant public health issue.

The aim of this rapid review is to highlight the evidence on effective interventions (and any relevant cost-effectiveness evidence) focusing on reducing the harms for children and young people who have been exposed to DVA.

Twenty-five studies were identified along with three guidance documents from the Welsh Government and the National Institute for Health and Care Excellence (NICE) in the UK. Twenty papers from nineteen studies reported the effectiveness of a wide range of interventions to support children and young people who have witnessed DVA. Most studies found meaningful differences in behaviour following an intervention. However, some studies did not find any differences between the intervention and control groups following an intervention to reduce the negative effects of witnessing DVA.

An included cost-effectiveness analysis suggested that for behavioural outcomes, a psychoeducational intervention delivered to parent and child in parallel is likely to be cost-effective among the interventions they compared. Two further full economic evaluation studies determined the cost-effectiveness of cognitive behavioural therapy interventions to support children and young people who have been exposed to DVA.

Policy and practice implications: Economic evaluations have found preliminary evidence that cognitive therapy is a cost-effective intervention to treat children and adolescents with PTSD. Future interventions should be co-produced with relevant stakeholders and patient and public members (including children and young people).

There is a need for larger, well conducted, pragmatic randomised controlled trials with longer follow-up periods.

**Funding statement:** The Bangor Institute for Medical and Health Research was funded for this work by the Health and Care Research Wales Evidence Centre, itself funded by Health and Care Research Wales on behalf of Welsh Government.

**EXECUTIVE SUMMARY:** *What is a Rapid Review?:* Our rapid reviews (RR) use a variation of the systematic review (SR) approach, abbreviating or omitting some components to generate the evidence to inform stakeholders promptly whilst maintaining attention to bias. They follow the methodological recommendations and minimum standards for conducting and reporting rapid reviews, including a structured protocol, systematic search, screening, data extraction, critical appraisal, and evidence synthesis to answer a specific question and identify key research gaps. They take 1- 2 months, depending on the breadth and complexity of the research topic/ question(s), extent of the evidence base, and type of analysis required for synthesis.

*Who is this summary for?:* This Rapid Review was conducted as part of the Health and Care Research Wales Evidence Centre Work Programme. The above question was suggested by members of the Communities and Tackling Poverty Group, Welsh Government, and a public representative for the Health and Care Research Wales Evidence Centre. The findings of the Review will inform the Violence against Women, Domestic Abuse and Sexual Violence (VAWDASV) National Partnership Board’s Children and Young Persons working group.

*Background / Aim of Rapid Review:* Children and young people witnessing domestic violence and abuse (DVA) can be affected negatively in terms of their psychological, emotional, and social development (An et al., 2017; Anderson, 2017). Adverse events in childhood (ACEs) are known to be harmful to a young person’s development and influence their life course (Campbell et al., 2016; Edwards, 2022; Lester et al., 2020), and therefore is a significant public health issue (Bellis et al., 2019). The long shadow cast by domestic abuse can influence the prospects and potential of individuals over the life course and beyond into future generations (Bellis et al., 2019; Edwards and McIntosh, 2019; Hardcastle et al., 2018; Hughes et al., 2021; Welsh Government, 2016a). The aim of this RR is to highlight the evidence on effective interventions (and any relevant cost-effectiveness evidence) focusing on reducing the harms for children and young people who have been exposed to DVA. The review question was: What is the effectiveness and cost-effectiveness of interventions in reducing the harms for children and young people who have been exposed to domestic violence or abuse? As part of an initial search for secondary evidence, a mixed method SR by Howarth et al (2016) was identified as a suitable basis upon which to build this RR. The Howarth et al (2016) SR was funded by the National Institute for Health Research (NIHR) and conducted in England (Howarth et al., 2016). This SR was specifically chosen because it included some economic evidence and reported evidence based on the type of domestic abuse interventions for children and young people. This RR builds upon Howarth et al (2016) by updating the evidence to include more recent studies.

*Key Findings:* Twenty-five studies were identified along with three guidance documents from the Welsh Government and the National Institute for Health and Care Excellence (NICE) in the UK.

*Effectiveness of interventions for those exposed to domestic violence and abuse:* Twenty peer-reviewed papers from nineteen studies reported the effectiveness of a wide range of interventions to support children and young people who have witnessed DVA. Interventions included advocacy services, psychoeducation, Cognitive Behaviour Therapy, play therapy and parenting skills training. Most studies found meaningful differences in behaviour following an intervention. However, some studies did not find any differences between the intervention and control groups following an intervention to reduce the negative effects of witnessing DVA.

*Cost-effectiveness of interventions for those exposed to domestic violence and abuse:* A cost-effectiveness analysis by Howarth et al (2016) suggested that for behavioural outcomes, a psychoeducational intervention delivered to parent and child in parallel is likely to be cost-effective among the interventions that they compared if willingness to pay was approximately £8000 (ICER = 3722 per Standard Mean Difference (SMD). Two further full economic evaluation studies determined the cost-effectiveness of CBT interventions to support children and young people who have been exposed to DVA (Aas et al., 2019; Shearer et al., 2018). Both the Aas et al (2019) and the Shearer et al (2018) interventions were deemed cost-effective alternatives relative to the control groups. Both studies were undertaken from health service and personal social services perspectives (although the authors of one of the studies did not explicitly state their perspective. Future studies may need to broaden their perspectives to consider wider costs to society (Edwards and McIntosh, 2019).

*Best quality evidence:* The best available economic evidence found in this the RR were the full economic evaluation studies that included both effectiveness and cost effectiveness elements (Aas et al., 2019; Shearer et al., 2018). The best quality evidence from the remaining 17 intervention studies reporting clinical effectiveness (which did not include full economic evaluations) were those that followed strict RCT methodology and subsequently scored well in our critical appraisal. All clinical effectiveness studies were deemed to be of moderate to high quality.

*Policy Implications:*

- Economic evaluations have found preliminary evidence that cognitive therapy is a cost-effective intervention to treat children and adolescents with PTSD.
- Future interventions should be co-produced with relevant stakeholders and patient and public members (including children and young people).

*Research Implications:*

- There is a need for larger, well conducted, pragmatic RCTs with longer follow-up periods. Robust full health economic evaluations for new and complex interventions in this area could include economic modelling once a solid evidence base exists.
- Information provided from the costing studies may be useful to inform future economic evaluations of interventions to support children and young people who have witnessed domestic abuse as they detail the key resources used for interventions.
- A wider societal perspective able to capture a broader set of costs and benefits, for example, possible parent productivity losses, warrants further consideration.

*Strength of Evidence:* All included studies were controlled trials, with most being RCTs. Certainty in the findings were moderate to low as most of the included studies had short time horizons and small sample sizes. Greater confidence in the findings would require a more robust evidence base.

## 1. BACKGROUND

### 1.1 Who is this review for?

This Rapid Review was conducted as part of the Health and Care Research Wales Evidence Centre Work Programme. The review question “What is the effectiveness and cost-effectiveness of interventions in reducing the harms for children and young people who have been exposed to domestic violence or abuse?” was suggested by members of the Communities and Tackling Poverty Group, Welsh Government and a public representative for the Health and Care Research Wales Evidence Centre. The findings from the proposed rapid review will inform the Violence against Women, Domestic Abuse and Sexual Violence (VAWDASV) National Partnership Board’s Children and Young Persons working group.

### 1.2 Background and purpose of this review

Domestic abuse relates to neglect, physical, emotional, and sexual abuse. Domestic abuse can impact the well-being of both the victim of abuse and those witnessing the abuse and can impact the survivors of abuse financially as well as physically or mentally. Housing issues also occur, and there are often difficulties with childcare (Welsh Government, 2022a). There are many longstanding impacts of domestic abuse that can affect the mental health and well-being of individuals throughout the life course (Hughes et al., 2021).

Children and young people witnessing domestic violence and abuse (DVA) can be affected negatively in terms of their psychological, emotional, and social development (An et al., 2017; Anderson, 2017). Since 5^th^ December 2022, children affected by domestic abuse are now automatically treated as victims regardless of whether or not they were present during violent incidents (The Crown Prosecution Service (CPS), 2023). The Crime Survey for England and Wales (CSEW) estimated that one in five adults aged 18 to 74 years experienced at least one form of child abuse, whether emotional abuse, physical abuse, sexual abuse, or witnessing DVA before the age of 16 years (8.5 million people) (Elkin, 2020). The National Society for the Prevention of Cruelty to Children (NSPCC)’s Speak out Stay safe (SOSS) programme for primary school children aims to increase children’s awareness and understanding of abuse and harm and enable them to seek help from a trusted adult (Stanley et al., 2021).

In the UK social restrictions caused by the COVID-19 pandemic led to children not being in places where they would normally be (Newbury et al., 2020). This meant that it was challenging for social workers, teachers and others to identify ‘at risk’ children and young people through typical safeguarding procedures in schools and extracurricular activities Regular support systems were difficult to access (National Institute for Health and Care Excellence, 2022). However, recent schemes such as ‘Ask me’ and ‘Live fear free helplines’ help those experiencing domestic violence (Davidge, 2020; Welsh Government, 2022b).

Children can thrive when they have a supportive adult that they can rely on to take them to school, monitor their educational attainment, attend parent-teacher meetings at school and ask about their interests and friends (Welsh Government, 2016b). This support can help a child to achieve their educational potential, have positive health and mental health outcomes, and make it more likely that they will develop good relationship and social skills (Clements and Fay-Hillier, 2019; Davies, 2019). In contrast, children and young people witnessing DVA in the home can be affected negatively in terms of their psychological, emotional, and social development, increasing disruptive behaviours, which may, in turn, cause difficulties for them at school (Hughes et al., 2017; Katz, 2016; Lloyd, 2018). Adverse Childhood Experience (ACE’s) are known to be harmful to a young person’s development and influence their life course (Campbell et al., 2016; Edwards, 2022; Lester et al., 2020), and therefore is a significant public health issue (Bellis et al., 2019). The aim of this RR is to highlight evidence of both the clinical effectiveness and cost-effectiveness of interventions focusing on reducing the harms for children and young people who have been exposed to domestic abuse. The review question is: What is the effectiveness and cost-effectiveness of interventions in reducing the harms for children and young people who have been exposed to domestic violence or abuse? This RR builds on the previous mixed method SR reported by Howarth et al (2016) (Howarth et al., 2016).

## 2. RESULTS

### 2.1 Overview of the Evidence Base

This RR was based on a SR by Howarth et al (2016), which was a mixed-method SR (Howarth et al., 2016). This 2023 RR search strategy was informed by Howarth et al (2016) but was amended to capture relevant cost-effectiveness evidence. This RR reports on the evidence identified from the Howarth et al (2016) SR and expands on this by presenting new evidence from 2015 to January 2023 (an updated search from the Howarth et al (2016) paper). Evidence from the SR conducted by Howarth and colleagues (2016) included trials and costing studies. The updated evidence in this RR included controlled trials and economic evaluations. The numbers of included studies are presented in the PRISMA diagram in Figure 6.1. The data extraction Tables and the quality appraisal tables can be viewed in Section 6 of this RR. Elements of domestic abuse guidance from the Welsh Government and the National Institute for Health and Care Excellence (NICE) in the UK were included for background information for the reader and are reported in Table 2.1. An evidence map of the literature is presented in Table 2.2, which categorises the evidence according to study intervention and study type.

#### 2.1.1 Guidance documents

Guidance documents include the 2016 and 2022 NICE guidance on domestic abuse (National Institute for Health and Care Excellence, 2022, 2016). There is also a relevant Welsh Government strategy document from 2016 (Welsh Government, 2016b). These guidance documents were not the correct study type to be eligible for data extraction. However, they offered important background and contextual information on the topic of domestic abuse, see Table 2.1.

**Table 2.1.** Elements of domestic abuse guidance in the UK

| Domestic abuse and violence guidance documents | Key elements of the guidance documents |
| --- | --- |
| National strategy on violence against women, domestic abuse and sexual violence: 2016 - 2021 (Welsh Government, 2016b). | The Welsh Government strategy was to:<br>• Increase awareness and challenge attitudes of violence against women, domestic abuse and sexual violence. |
|  | • Increase awareness in children and young people of the importance of safe, equal and healthy relationships.<br>• Increase focus on holding perpetrators to account and provide opportunities to change their behaviour.<br>• Make early intervention and prevention a priority.<br>• Ensure relevant professionals are trained to provide effective, timely and appropriate responses to victims and survivors.<br>• Provide victims with equal access to appropriately resourced, high quality, needs led, strength based, gender responsive services across Wales. |
| Domestic abuse quality standards (National Institute for Health and Care Excellence, 2016) | The quality standard is expected to contribute to improvements in the following outcomes:<br>• Harm from domestic violence and abuse<br>• Mortality from domestic violence and abuse<br>• Emergency attendances for domestic violence and abuse<br>• Quality of life<br>• Personal safety<br>• Duration of domestic violence and abuse<br>• Re-occurrence of domestic violence and abuse. |
| Recognising and responding to domestic violence and abuse (National Institute for Health and Care Excellence, 2022) | This guidance for social workers suggested that social workers should know how to respond to domestic violence and abuse and how to refer to specialist services for additional support. Services include:<br>• Refuges<br>• Advocacy<br>• Floating support<br>• Outreach support<br>• Specialist support for perpetrators<br>• Tailored interventions<br>• Housing workers<br>• Independent domestic violence advisors (IDVA)<br>• Multi-Agency Risk Assessment Conference (MARAC) for people at high risk.<br>This guidance also stated that social workers should be aware that some people may find domestic violence and abuse services more difficult to access or use. This may include older people, men, people with disabilities, people from black and minority ethnic groups, and lesbian, gay, bisexual, or trans people. It may also include people who are not entitled to benefits, housing, and other public services, usually due to their immigration status. Advocacy and additional support may help with this. |

**Table 2.2.**
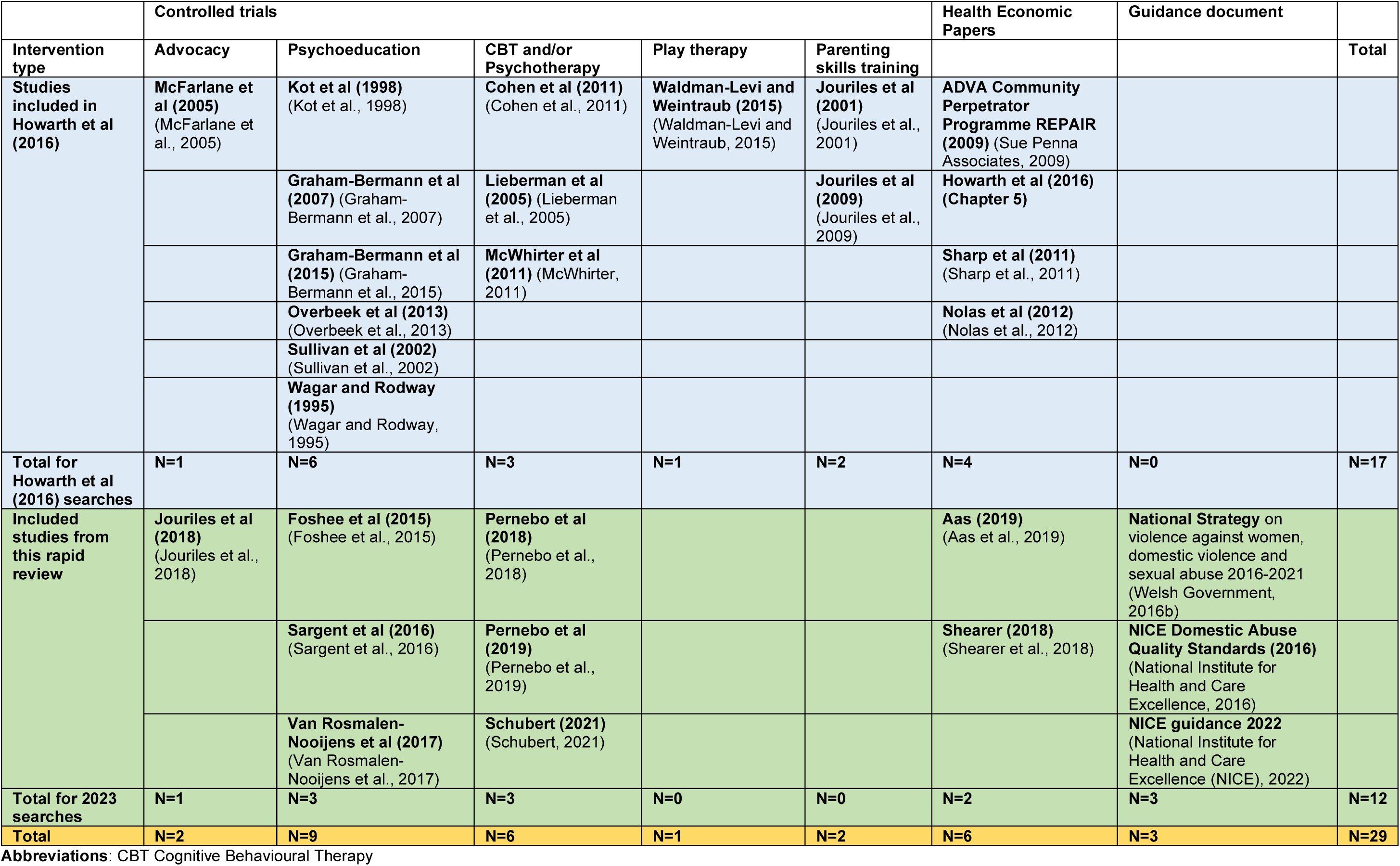
**Map of included domestic violence and abuse studies according to type of intervention (and guidance documents)**

### 2.2 Clinical effectiveness of interventions targeting children and young people who have witnessed domestic abuse

In this RR, twenty peer-reviewed papers describing nineteen studies were included. This included the sixteen studies identified by Howarth et al (2016), (the Howarth et al (2016 cost-effectiveness analysis included eight of these and all were from the USA) and twelve recent studies from the new searches. All the studies investigated the clinical effectiveness of interventions to reduce the impact of witnessing domestic abuse. Two papers included in this RR investigated the cost-effectiveness of their respective intervention programmes.

Most included studies were from the United States of America (USA) (n=13) (Cohen et al., 2011; Foshee et al., 2015; Graham-Bermann et al., 2007, 2015; Jouriles et al., 2018, 2009, 2001; Lieberman et al., 2005; McFarlane et al., 2005; McWhirter, 2011; Sargent et al., 2016; Schubert, 2021; Sullivan et al., 2002), two papers were from Canada (n=2) (Kot et al., 1998; Wagar and Rodway, 1995), two were from the Netherlands (n=2) (Overbeek et al., 2013; Van Rosmalen-Nooijens et al., 2017), two were from Sweden (n=2) (Pernebo et al., 2019, 2018), and one was from Israel (n=1) (Waldman-Levi and Weintraub, 2015).

The clinical effectiveness studies are described according to type of intervention below, and the health economic studies are presented at the end of this section. Identified Interventions included advocacy services, psychoeducation, Cognitive Behaviour Therapy (CBT), play therapy and parenting skills training.

#### 2.2.1 Advocacy Services

Two of the included studies considered the effectiveness of Advocacy Services in the USA (Jouriles et al., 2018; McFarlane et al., 2005). The first of these studies was an advocacy support-based RCT nurse case-management intervention study for children of abused mothers (McFarlane et al., 2005). In this study, two interventions were compared, using analysis of variance (ANOVA) wallet card with no support and wallet card with support.

There was no difference in outcomes between the two intervention groups. The child behaviour scores were improved for both groups. All children’s scores improved on the Child Behaviour Checklist (CBCL) over the treatment period. There was a significant main effect of time of administration (F (8,94) = 16.18, p< .001 (and) CBCL scores (F(8,121) = 11.08, p < .001) for children 18 months to 5 years of age, and youth, 6 to 18 years of age). The authors concluded that taking away the secrecy and privatisation of domestic violence may have interrupted and prevented the reoccurrence of domestic abuse, which resulted in more positive outcomes for the children. Although the McFarlane (2005) study had an intervention and control group, there was incomplete outcome data, and this influenced the validity of the study findings.

The other advocacy study by Jouriles et al (2018) was a secondary analysis of an RCT using multi-level modelling methodology to investigate the effectiveness of advocacy support for abused mothers and their children aged between 4 and 9 years old (n=66). They found that Project Support reduced the extent of partner–child contact. In addition, within-subject changes in contact over time were associated with the conduct problems of girls but not boys, and it partially mediated the effects of Project Support on girls’ conduct problems.

Multilevel modelling analysis results indicated a Deviation in Contact × Child Sex interaction effect, b = –.16, t(241) = −3.45, p< .001, d = .44; positive deviations in contact related positively to conduct problems for girls (the “b” path in the mediation model), b =.11, t(239) = 3.00, p< .005, d = .39,2 but not for boys. Higher average levels of contact over time child and partner–child aggression helped explain the effects of contact on children’s conduct problems (Jouriles et al., 2018). The main limitation of this study is that most of the outcomes were observational, mother reported outcomes. Furthermore, internal consistency of some of the included measures was low, affecting measurement validity.

#### 2.2.2 Psychoeducation

Nine included studies focused on a psychoeducation intervention (Foshee et al., 2015; Graham-Bermann et al., 2007, 2015; Kot et al., 1998; Overbeek et al., 2013; Sargent et al., 2016; Sullivan et al., 2002; Van Rosmalen-Nooijens et al., 2017; Wagar and Rodway, 1995). Five of the psychoeducation studies were conducted in the USA, two of the studies were conducted in Canada, and two were conducted in The Netherlands.

Three of the included studies focused on psychoeducation interventions and behavioural outcomes (Graham-Bermann et al., 2007; Kot et al., 1998; Wagar and Rodway, 1995).

Wagar and Rodway (1995) conducted an RCT in Canada with a group treatment including education and knowledge about how to keep safe, and a control group. ANOVAs were conducted within this study, which found that children in the intervention group had improved scores on attitudes and responses to anger (F = 8.23, p < .01), and sense of responsibility for their parents and the violence they experienced (F =4.50, p < .05). Knowledge of safety and support did not differ between the treatment and control groups (p > .05) (Wagar and Rodway, 1995). Although this was a study using RCT methodology, the reporting was not clear as to how the children and young people were randomised and no objective outcomes were included.

Child behaviour was also the focus of the RCT study by Kot (Kot et al., 1998). Kot et al (1998) investigated child-centred play therapy with child witnesses of domestic abuse in Canada and found that play therapy within a period of three weeks was effective, and child behaviours and self-concepts improved. Analysis of Covariance (ANCOVA) analyses were conducted, and it was found that children in the experimental group scored significantly higher than the children in the control group on self-concept as measured by the Joseph Pre-School and Primary Self-Concept Screening Test (JPSST): F (1, 19) = 48.96, p< .001. Children in the experimental group scored significantly higher than children in the control group in the Children’s Play Session Behaviour Rating Scale (CPSBRS) areas of Physical Proximity, F (1, 19) = 13.561, p<.01; and Play Themes, F (1,19) = 12.18, p < .01. There was also a significant (p < .05) (no confidence intervals were reported) reduction in externalising behaviour problems for the experimental group at post-test. The mothers of children in the experimental group perceived their children as less aggressive and as manifesting fewer delinquent behaviours such as lying, cheating, and swearing. This decrease in behaviour problems was perceived by the mothers to be particularly noteworthy. Caution should be taken when interpreting these findings as no formal randomisation was conducted, no blinding was conducted, and no objective measures were used.

Graham-Bermann et al (2007) conducted an RCT to investigate the effectiveness of a psychoeducational intervention to reduce conduct problems among children of parents experiencing intimate partner violence (IPV) in the USA. During the intervention period (10 weeks in length), child conduct problems decreased in the Project Support group, b=-.53, t(64)=-7.13,p<.001, as well as in the comparison group (waitlist control), b=-.30,t(64)=- 5.16,p<.01. However, they decreased more rapidly in the Project Support group than in the comparison group, b =.23, t(64) = 2.78, p < .02. Oppositional child behaviour decreased more slowly than other measures of child conduct problems, b = .39, t(332) = 492, p < .001 (Graham-Bermann et al., 2007). As with Kot (1998), caution should be taken when interpreting these findings as no formal randomisation was conducted, and no objective measures were used.

Two of the included psychoeducation studies focused on mental health disorders such as depression, anxiety and post-traumatic stress disorder (PTSD) (Overbeek et al., 2013; Van Rosmalen-Nooijens et al., 2017). Overbeek et al (2013) investigated PTSD of children aged 6 to 12 years in The Netherlands. They found that the 9-week group therapy intervention was effective in improving the emotional and coping skills of children who had experienced post-traumatic stress as a consequence of IPV (Overbeek et al., 2013). However, only subjective outcomes were used for the analyses.

Another RCT conducted in The Netherlands included seventeen participants (intervention: n=8, control: n=9) (Van Rosmalen-Nooijens et al., 2017). Mixed model analysis showed significant differences between the intervention and control groups on the Symptom Checklist Depression (SCL-90 DEP) (P < .05) and Symptom Checklist Anxiety (SCL-90 ANX) (P < .05) subscales between 6 and 12 weeks after participation started. However, a Univariate Analysis of Variance (UNIANOVA) showed no significant differences between the intervention and control group at the final follow-up. There was no significant difference between the intervention group and the control group at the final follow-up on the Impact of Event Scale (IES), P > 0.1. In terms of additional findings, all participants involved with the internet-based intervention felt safe (Van Rosmalen-Nooijens et al., 2017). The main limitation of this study was that no male participants completed their participation. Therefore, the results are only relevant for females.

One of the included papers focused on child internalised problems as the main outcomes of their psychoeducational intervention (Graham-Bermann et al., 2015). A second RCT by Graham-Bermann et al (2015) was conducted in community settings in the USA with children of abused mothers. The hypothesis that children in the intervention condition would show significant improvement in internalizing symptoms, relative to those in the no treatment comparison group was partially supported. The interaction of treatment and time at follow-up was statistically significant, indicating that for girls in the treatment group, there was a statistically significant decrease in internalising problems at the 8 month follow up point. The time trajectory for boys in the treatment group was computed by creating a linear combination of the appropriate main effects and interaction terms and was not statistically significant. Effect sizes (Cohen’s d) from baseline to follow-up were .18 for the treatment group and .15 for the comparison group. Effect sizes of change from Time 2 to Time 3 were small (.01 for the treatment and -.07 for the comparison group). As with the previous study conducted by these authors, this study also had a high risk of bias, failing on selection, performance and attrition (Graham-Bermann et al., 2015).

Dating abuse outcomes were investigated in an RCT conducted in the USA (Foshee et al., 2015). This study investigated the effectiveness of the ‘Mothers and Teens for Safe Dates’ (MTSD) program, which was found to have significant program effects on the perpetration of cyber abuse (p < .0.05), but not for adolescents who had average or low exposure to DVA (p > 0.1). There were no program effects on the perpetration of physical or sexual dating abuse (Foshee et al., 2015). The outcome measures used by Foshee et al (2015) were new, with no psychometric testing conducted on them, which could affect internal validity and impact generalisability.

One study focussed on global self-worth as a main outcome of a psychoeducational intervention (Sullivan et al., 2002). The RCT conducted in the USA found that the strength-based intervention for women and their children who have experienced DVA was moderately effective. Children in the experimental condition reported significantly higher self-competence in several domains (e.g., global self-worth, physical appearance, and athletic sub-scale) compared to children in the control group (p <.05). The authors noted that the measures used to measure intervention effectiveness were not sensitive enough (Sullivan et al., 2002). The main limitation of this study is the lack of robust outcome measures.

One study focussed on knowledge of DVA (Sargent et al., 2016). This RCT conducted in the USA investigated the effects of an online educational program in increasing knowledge about children’s exposure to DVA. The community group and a college student group improved their knowledge of DVA and their self-efficacy in helping children who have experienced DVA after being involved in the Change A Life intervention (P < .001). The community control group, who completed an on-line programme from the Alzheimer’s Association, did not show an improvement in their knowledge of DVA (P > 0.1). Neither participant gender nor prior exposure to DVA moderated the intervention effects (Sargent et al., 2016). Limitations of this study by Sargent et al (2016) included a very short follow-up period (one week) and no objective outcome measures were used (the evaluation was limited to participants’ self-reports of their knowledge of DVA.

#### 2.2.3 Cognitive Behavioural Therapy and/or Psychotherapy

Six studies focussed on CBT and/or Psychotherapy. Three were identified in the Howarth et al (2016) (Howarth et al., 2016) SR, (Cohen et al., 2011; Lieberman et al., 2005; McWhirter, 2011), and three more were identified in the search for this RR (Pernebo et al., 2019, 2018; Schubert, 2021). The studies were from the USA and Sweden and were conducted between 2005 and 2021.

In 2005, Lieberman et al (Lieberman et al., 2005) investigated the effectiveness of child-parent psychotherapy versus usual care over 50 weeks in community settings in the USA. Analyses of CBCL total scores showed a significant group x time interaction, F1,61 = 5.77, p < .05, d = 0.24, with follow-up analyses revealing that only the CPP group evidenced significant intake-post-test reductions: t (34) = 2.86, p < .01. To examine whether the error was introduced because some children completed the CBCL 2-3 at intake and the CBCL 4- 18 at post-test, analyses were repeated with only the children who completed the CBCL 4-18 at intake and post-test. These analyses also resulted in a significant interaction effect (F1,31 = 4.72, p < .05, d = 0.64), with follow-up analyses confirming that only the CPP group showed significant reductions in behaviour problems (CPP: intake mean = 60.32, SD = 9.00; post-test mean = 54.16, SD = 8.71, t (18) = 3.10, p < .01; comparison: intake mean = 58.86, SD = 8.82; post-test mean = 59.64, SD = 13.11). The overall findings highlight the importance of including the mother as an integral partner in the treatment of pre-schoolers’ traumatic stress symptoms (Lieberman et al., 2005). It was inferred that there were 42 children in the treatment group and 31 children in the comparison group and a 13% dropout rate was observed. However, the reasons for attrition were not stated clearly, suggesting that the reasons for dropout were unclear. The reasons may include, but are not limited to, a bad experience or dissatisfaction with the intervention.

In another study from the USA, Cohen et al (2011) conducted a RCT in community settings to investigate the effectiveness of an 8-week CBT intervention for (n=124) 7 to 14-year-olds. They found that the children in the intervention group made greater improvements than children in the usual care groups. Children completing Trauma Focused-Cognitive Behavioural Therapy (TF-CBT) had significantly greater improvement than children completing Child Centred Therapy (CCT) in Kiddie Schedule for Affective Disorders and Schizophrenia, Present and Lifetime Version (K-SADS-PL) total score (1.67; −0.08 to 3.4) and Reaction Index score (−7.58; −0.79 to −14.38) and in K-SADS-PL hyperarousal score (−0.81; −0.03 to −1.59) and anxiety score (−7.36; −1.06 to −13.67). Following the intervention, children could feel safer in the face of ongoing danger; for example, differentiating between real and generalized fears, learning safety and relaxation strategies, and talking directly to the mother about IPV experiences. The main limitation of the Cohen et al (2011) study was that they did not use objective outcomes and there was incomplete outcome data, limiting the generalisability of the findings.

In another RCT investigating the effectiveness of emotion focussed group therapy and goal focussed group therapy conducted in 2011 (McWhirter, 2011), temporarily homeless families in the USA (mothers and their children) were asked to complete emotional and psychosocial measures. The child participants fundamentally responded in positive ways to both interventions (emotion and goal focussed group therapy). The results of this study are congruent with studies that demonstrate the efficacy of family-based interventions involving the child with his or her mother following IPV. Multicomponent approaches involving mothers and children successfully improved attitudes about violence and reduced aggression among children exposed to DVA (McWhirter, 2011). This study by McWhirter (2011) had very few participants and had a very short follow-up period, therefore the results should be viewed with caution.

A trial conducted in 2021 in the USA compared a ‘Child Witness to Domestic Violence programme’ (CWDV) for mothers and children with a control group (Schubert, 2021). As a quasi-experimental design was used with different time periods for the intervention and the control, the risk of bias is high. However, it was found that children who participated in CWDV demonstrated less hyperactivity, fewer negative emotional symptoms, and fewer total behavioural difficulties than their peers who did not participate in CWDV. Specifically, multiple regression analyses indicated that condition (intervention vs. control) was a significant predictor of child hyperactivity (B = –.85, p = <.05; mean group difference at post-test = 0.63 out of 10), negative emotional symptoms (B = –1.14, p = < .01; mean group difference at post-test = 1.22 out of 10), and total behavioural difficulties (B = –2.48, p = .02; mean group difference at post-test = 2.23 out of 40) (Schubert, 2021). Limitations of the Schubert (2021) study included a reliance on maternal reporting.

In Sweden, two publications by Pernebo et al (Pernebo et al., 2019, 2018) were published, reporting on psychotherapy and psycho-educational interventions. Pernebo et al (2018) conducted a study that compared psychotherapy (n=19) and psycho-educational interventions (n=31) for children and young people exposed to IPV. Both interventions were 12-15 weeks of 90-minute sessions per week. The mothers in the Community Based Intervention (CBI) reported a significant reduction in their child’s emotional symptoms following the interventions. Strengths and Difficulties Questionnaire (SDQ-P); d=0.34), in total post-traumatic stress Trauma Symptom Checklist for Young Children (TSCYC); d=0.35), and in intrusive symptoms (TSCYC; d=0.40). Mothers in the CBI additionally reported a significant decrease in impact scores (SDQ-P; d=0.62). The mothers in the CAMHS intervention (CAMHSI) reported significant reductions in their child’s symptoms in several areas: overall mental health symptoms (SDQ-P; d=0.67), emotional symptoms (SDQ-P; d=0.73), hyperactive symptoms (SDQ-P; d=0.46), impact score (SDQ-P; d=0.68), emotionality Emotion Questionnaire for Parents (EQ-P); d=0.57), and (TSCYC) symptoms of anger (d=0.65), arousal (d=0.66), and dissociation (d=0.76). The mothers reported large effects in the CAMHSI for a decrease in depressive symptoms (TSCYC; d=0.99) and an increased capacity for emotion regulation (EQ-P; d=0.85) (Pernebo et al., 2018). The results of the study indicate that the psychotherapeutic intervention was somewhat more effective than the psychoeducation intervention in reducing child symptoms in the aftermath of IPV. The lack of a control group limits the conclusions that can be drawn from the results. The Pernebo et al (2019) paper reported on the same study but expanded upon it to assess the long-term impacts of the two interventions at 6 and 12 months (Pernebo et al., 2019).

Significant improvements in children’s symptoms of general psychological health and trauma symptoms were reported by mothers from pre-intervention to the follow-up assessments (p = .004– .044; d = 0.29–0.67). However, there was a reduction in exposure to violence as self-reported by mothers. The small sample size in the research by Pernebo et al (2018, 2019) limited the authors ability to conduct subgroup analysis (different ages, ethnicities, experiences, and living conditions), which was one of the main limitations of the study.

#### 2.2.4 Play therapy

Howarth et al (2016) identified one study focussing directly on occupational performance and play therapy (Waldman-Levi and Weintraub, 2015). No new studies (post-2015) focussing on play therapy were identified for this RR. Waldman-Levi et al (2015) (Waldman-Levi and Weintraub, 2015) conducted a pre-test/post-test two-group control study design in Israel with twenty mother-child dyads (children aged between 1 and 6 years old). The intervention aimed to improve occupational performance and there was also a playroom program which acted as the control arm. After the intervention, mother–child interaction was significantly better in the Family Intervention for Improving Occupational Performance (FI–OP) group than in the playroom group. The Mann–Whitney U test used to compare the study groups’ difference scores (pre-test vs. post-test) with respect to children’s playfulness (ToP) revealed no significant difference between the FI–OP and playroom groups. The results of this study indicate that children in the FI–OP program significantly improved their play skills compared with the playroom group, in which no significant improvement was noted. The creation of a safe space during the intervention may facilitate mother-child interaction. However, there was no follow-up phase in this study, limiting the overall findings of the eight-week intervention.

#### 2.2.5 Parenting skills training

Two papers included in the Howarth et al (2016) SR focussed on parenting skills training (Jouriles et al., 2009, 2001). Jouriles et al (2001) investigated conducted an RCT intervention focussing on teaching mother’s child management skills compared to usual care of children with a conduct disorder. The results evaluating the children over time showed a significant interaction (p<.01) on the CBCL checklist. The children in the treatment condition improved at a faster rate (slope -3.53) than the children in the control condition (slope = -.95). Children were assessed 5 times in all. By assessment three, there was no difference between the groups. The study by Jouriles et al (2001) did not use objective measures, thus limiting the generalisability of the findings.

In 2009, the same authors published further work with mothers and children recruited from shelters, this time with a larger sample (Jouriles et al., 2009). During the intervention period, child conduct problems decreased in the Project support group (P <.001) as well as in the comparison group (P <.01). However, they decreased more rapidly in the Project Support group than in the comparison group (P <.01). For the follow-up period, conduct problems continued to decrease in the Project support group (P < .005) but not in the comparison group (P > .05) . The authors noted that the effectiveness of the Project support intervention is dependent on the mother’s general well-being and parenting skills (Jouriles et al., 2009). The Jouriles et al., (2009) study used random allocation, but did not use objective measures and therefore caution should be taken to when interpreting the results.

#### 2.2.6 Bottom line results for the clinical effectiveness interventions

This RR identified twenty peer-reviewed papers from nineteen studies reporting on the effectiveness of a wide range of interventions to support children and young people who have witnessed DVA. The papers were deemed to be of moderate to high quality following critical appraisal. Most of the studies looked at short-term outcomes rather than longer-term outcomes. There was a lack of studies including the outcomes of educational attainment, school/college attendance and school/college functioning.

Two studies assessed the effectiveness of advocacy services. One study reported no difference in effectiveness between a nurse case management intervention and the control group (McFarlane et al., 2005). In contrast, the other advocacy services study of a secondary analysis of data from a RCT found that participation in the intervention resulted in favourable outcomes such as the decreased frequency of child contact with their mother’s violent partner (Jouriles et al., 2016).

Nine studies reported on the effectiveness of psychoeducation interventions. Three studies reported a reduction in behavioural problems following the intervention (Graham-Bermann et al., 2007; Kot et al., 1998; Wagar and Rodway, 1995). Two studies reported on emotional outcomes after post-traumatic stress as a consequence of DVA (with one study showing an effective outcome post intervention (Overbeek et al., 2013) and the other not (Van Rosmalen-Nooijens et al., 2017)). One psychoeducational study found that an intervention helped girls with not internalising problems at the follow-up, but not boys (Graham-Bermann et al., 2015). One study found that online dating abuse was reduced as a consequence of an intervention in dating after exposure to DVA (Foshee et al., 2015). One study found that self-worth was significantly higher for those in the intervention group compared to those in the control group (Sullivan et al., 2002), and one study found that self-efficacy increased in those in the Change a Life intervention compared to those in the control group (Sargent et al., 2016).

Six studies measured the effectiveness of CBT and/or psychotherapy. Two of the studies highlighted the importance of including the mother in the treatment of children with traumatic stress symptoms due to exposure to DVA (Lieberman et al., 2005; McWhirter, 2011). One study found that trauma focused CBT was more effective than child-centred therapy only following IPV experiences (Cohen et al., 2011). Two papers found that psychotherapy was more effective in reducing child symptoms in the aftermath of IPV than psychoeducation (Pernebo et al., 2019, 2018), and one study found that children who had taken part in the CWDV programme demonstrated less hyperactivity and fewer emotional symptoms and fewer behavioural difficulties than those not participating in CWDV (Schubert, 2021).

One study reported favourable findings for a play therapy intervention, which showed significantly improved mother-child interaction in the intervention group in comparison to the control group (Waldman-Levi and Weintraub, 2015).

Of the two studies reporting on the effectiveness of parenting skills training, both reported greater reductions in conduct problems for children in the Project Support intervention group compared with those in the comparison group (Jouriles et al., 2009; 2001). See Summary of clinical effectiveness studies in Table 2.3.

**Table 2.3.** **Summary of clinical effectiveness studies**

| Author and date | Type of intervention | Primary outcome (Effective Yes (Y) /No (N)) |
| --- | --- | --- |
| Jouriles et al (2018) | Advocacy | Y |
| McFarlane et al (2005) | Advocacy | N |
| Cohen et al (2011) | CBT/Psychotherapy | Y |
| Lieberman et al (2005) | CBT/Psychotherapy | Y |
| McWhirter et al (2011) | CBT/Psychotherapy | Y |
| Pernebo et al (2018) | CBT/Psychotherapy | Y |
| Pernebo et al (2019) | CBT/Psychotherapy | Y |
| Schubert (2021) | CBT/Psychotherapy | Y |
| Jouriles et al (2001) | Parenting skills training | Y |
| Jouriles et al (2009) | Parenting skills training | Y |
| Waldman-Levi and Weintraub (2015) | Playtherapy | Y |
| Foshee et al (2015) | Psychoeducation | Y |
| Graham-Bermann et al (2007) | Psychoeducation | Y |
| Graham-Bermann et al (2015) | Psychoeducation | Y (girls not boys) |
| Kot et al (1998) | Psychoeducation | Y |
| Overbeek et al (2013) | Psychoeducation | Y |
| Sargent et al (2016) | Psychoeducation | Y |
| Sullivan et al (2002) | Psychoeducation | Y |
| Van Rosmalen-Nooijens et al (2017) | Psychoeducation | N |
| Wagar and Rodway (1995) | Psychoeducation | Y |

### 2.3 Economic evidence of interventions targeting children and young people who have witnessed domestic abuse

This RR aimed to build upon the Howarth et al (2016) SR and identify recent evidence on the cost-effectiveness of interventions comparing both costs and outcomes/benefits. Cost-effectiveness analysis by Howarth et al (2016) suggested that for behavioural outcomes, a psychoeducational intervention delivered to parent and child in parallel is likely to be cost-effective among the interventions that they compared if willingness to pay was approximately £8000 (ICER = 3722 per Standard Mean Difference (SMD). or mental health outcomes, it is very likely that a psychoeducational intervention delivered to the child would be cost-effective. If willingness to pay per SMD in mental health outcomes is high (ICER > £22,575/SMD), cognitive–behavioural therapy (delivered to the parent, child and dyad) may be equally cost-effective. Costs were estimated based on the reported description of the interventions in the study publications. They found much heterogeneity in the data that reflected the complex nature of these interventions. Training costs were not included in the analysis before this would be a one-off cost and not an annual cost for rolling interventions. The variety of intervention venues were not costed either and neither were ongoing supervision costs. Therefore, there was a large degree in uncertainly about the intervention costs, which is a limitation of this cost-effectiveness analysis. Howarth et al (2016) noted that their analyses were intended to be ‘hypothesis-generating’ to inform the future design of research studies, rather than robust estimates of effectiveness and cost-effectiveness.

Therefore, their tentative conclusions concerning which interventions to pursue in future research studies should be treated with caution.

The updated search of the literature identified two full economic evaluations (Aas et al., 2019; Shearer et al., 2018). The findings of this RR identified a clear lack of cost-effectiveness evidence in this area. Consequently, we broadened our scope to include costing only studies from the Howarth et al (2016) SR (Nolas et al., 2012; Sharp et al., 2011; Sue Penna Associates, 2009), which may be helpful to inform future economic evaluations in this area.

#### 2.3.1 Cost-effectiveness studies

Two recent full economic evaluations were included in this review, one from England (appraised to be moderate quality) and one from Finland (appraised as high quality). In 2018, Shearer published a paper which investigated the cost-effectiveness of a CBT intervention in the East of England, UK (Shearer et al., 2018). This decision analytic model-based cost-utility analysis (a form of cost-effectiveness analysis) was undertaken from an NHS and Personal Social Services perspective. The intervention group received ten weekly sessions of Cognitive Therapy for PTSD (CT-PTSD) delivered by a trained clinical psychologist. The waitlist control group received usual care provided by the NHS. The primary economic outcome was quality-adjusted life years (QALYs) which were mapped from the parent-completed Strengths and Difficulties Questionnaire (Furber et al., 2014; Furber and Segal, 2015). Costs were presented in British Pound Sterling (GBP) for cost year 2014. Costs and outcomes were discounted at the rate of 3.5% after the first year to reflect time preferences. This study provided preliminary evidence for the cost-effectiveness of CT- PTSD for children and young people with PTSD who had been exposed to at least a single traumatic event in the previous 2-6 months. The results of the model-based cost-utility analysis found that the ICER, after a three-year discounting rate for the CT-PTSD intervention compared to usual care, generated a cost per QALY of £2,205. It was concluded that the intervention was cost-effective in the UK at the current NICE threshold of £20,000 to £30,000 per QALY (Shearer et al., 2018). The treatment effect was significant, and patients in the intervention group gained more QALYs than untreated ones (2.370, 2.324), with a difference of 0.0577 between groups. Using compete case data only, probabilistic sensitivity analysis was conducted to vary the baseline assumptions. The cost-effectiveness acceptability curve (CEAC) demonstrated that the probability of CT-PTSD being a cost-effective alternative relative to usual care was between 69% and 75% at the current NICE cost-effectiveness threshold (Shearer et al., 2018). The main limitations of this Shearer et al (2018) study were the small numbers in the CT-PTSD intervention group (n=12), the short follow-up period of only 11 weeks and many assumptions made in the extrapolating to the model time horizon.

In 2019 Aas et al., (2019) conducted a RCT with an embedded economic evaluation to assess the effectiveness and cost-effectiveness of a CBT intervention focused on trauma in Finland (Aas et al., 2019). This trauma included: physical abuse within the family, witnessing physical violence within the family and sexual abuse outside the family. This economic evaluation was a cost-utility analysis and used the 16D (16-Dimension Quality of Life measure for adolescents) (Apajasaio and Hoimberg, 1996) as a measure of health-related quality of life (HRQoL) to derive QALYs. The authors did not state their perspective of analysis within the paper. Costs were presented in Norwegian Krone (NOK) at 2018 prices. There was no significant difference in total minutes of therapy and costs between the intervention (TF-CBT) and control (usual treatment) groups. HRQoL increased in both the intervention and control groups, but there were no significant differences in QALYs between the groups (1.573 for TF-CBT and 1.536 for TAU; p = .281). The CEAC demonstrated that it is very likely (96%) that TF-CBT is a cost-saving alternative and that the use of other services will decline, such as welfare services, medication, and school nurses. The authors concluded that TB-CBT should be advocated as the standard treatment for children and young people presenting with PTSD but also acknowledged a high drop-out rate, which was a major limitation in this study. See Table 2.4 for the ICERs for Aas et al (2019) and Shearer et al (2018).

**Table 2.4.** **Summary of Cost-effectiveness studies**

| Study | Mean cost | QALYs | Incremental |  | ICER |
| --- | --- | --- | --- | --- | --- |
|  |  |  | Cost | QALYs |  |
| **Aas et al (2019)** |  |  |  |  |  |
| TF-CBT | 5,935 NOK<br>(£463.52) | 1.573 |  |  |  |
| Usual care | 7,430 NOK<br>(£580.29) | 1.533 | -1,495 | 0.040 | -37,375<br>NOK<br>(dominant)<br>(£2,918.99) |
| **Shearer et al (2018)** |  |  |  |  |  |
| **3 year discounting** |  |  |  |  |  |
| CT-PTSD | £4,865 | 2.370 |  |  |  |
| Usual care | £4,768 | 2.324 | £97 | 0.0577 | £2,205 |

#### 2.3.2 Cost analysis studies

Three cost analysis studies were reported in the SR conducted by Howarth and colleagues (Howarth et al., 2016). Firstly, the REPAIR perpetrator programme established in three areas of Devon that focused on motivation, responsibility, safety, and acknowledgement for women, men and children was assessed. The women’s services were based on individual needs where a woman’s support worker provided advocacy, and practical and psychological support. The children’s groups focused on safety, risk assessment, the development of resilience, appropriate coping strategies and support networks and processing difficult feelings, along with the element of liaising closely with the school and especially with the classroom teacher. The majority of the 20 children of fathers on the perpetrator programme (where the father was the perpetrator) demonstrated decreased anxiety, stress and anger and an improved relationship with mothers and peers, as reported by the mothers (Sue Penna Associates, 2009). The net benefit per annum was £158,890 (this was the estimated difference between the total cost to society of £345,280 and the total cost of the REPAIR programme, which was £186,390 per annum). The REPAIR programme would serve 24 families per year (including mothers, fathers and children) (Sue Penna Associates, 2009).

Another study conducted in the UK by Sharp et al (2011) found that the overall costs of the pilot twelve-week psychoeducation programme at three sites in Scotland was £837,303 over a three-year period (Sharp et al., 2011). The largest driver of these costs was intervention coordinator salary of £99,820. The authors acknowledged that the available data included in the analysis was limited to short-term service delivery costs. To fully establish whether the psychoeducation programme provides good value for money, the authors recommended further assessment to consider all relevant costs, including both direct and indirect costs, short and longer-term costs and the potential for savings (which may not be directly or immediately quantifiable).

The third costing study identified in the SR conducted by Howarth and colleagues was an evaluation of a community group programme for children and young people in England (Nolas et al., 2012). The group psychoeducation programme provided support to children and young people to help them process their experiences of witnessing DV. Estimated delivery costs for running the twelve-week programme with seven children/young people was a little over £1,300 in 2012 (this would be £1,725 in December 2022 (Bank of England Inflation Calculator). Sex balance in the children’s groups was important to children and they valued attending separate groups from their siblings. For a minority of children, the timings of the groups had been inconvenient, as they missed out on school-curricular and extracurricular activities (Nolas et al., 2012).

#### 2.3.3 Bottom line results for the health economics studies

Despite being sought for inclusion, only two full economic evaluations assessing the cost-effectiveness of interventions to support children and young people who have been exposed to domestic abuse were identified (Aas et al., 2019; Shearer et al., 2018). Both studies were deemed as cost-effective alternatives relative to the control groups. Future studies in this area may wish to broaden their perspective to consider wider costs to society as ACEs are known to be harmful to the development of children and young people and influence their life course), and therefore is a significant public health issue (Bellis et al., 2019; Campbell et al., 2016; Edwards, 2022).

The three cost analysis studies presented intervention programme costs for various interventions to improve the outcomes for children and young people who had witnessed domestic abuse. Information provided from the costing studies may be useful to inform future economic evaluations of interventions to support children and young people who have witnessed DVA. These three cost analysis studies could not be quality appraised appropriately due to the lack of a standardised cost analysis quality appraisal checklist/tool (Xu et al., 2021). These cost analysis studies consistently failed to meet the CHEERS checklist criteria for economic modelling, economic assumptions, modelling parameters, and outcome measures. However, the full economic evaluations largely met these criteria, resulting in higher critical appraisal judgements for the full economic evaluations over the partial economic evaluations.

## 3. DISCUSSION

### 3.1 Summary of the findings

This RR provides evidence of the effectiveness of interventions aimed at children and adolescents exposed to domestic abuse. This was an important question to investigate as there is a long shadow cast by domestic abuse that influences individuals’ prospects and potential over the life course (Bellis et al., 2019; Edwards and McIntosh, 2019; Hardcastle et al., 2018; Hughes et al., 2021). Future generations may also be affected by the legacy of the previous generations (Welsh Government, 2016a).

The included studies represent an international body of evidence. In summary, a total of twenty-four primary studies using controlled trial methodology measured the effectiveness of interventions to improve the outcomes for children who had witnessed domestic abuse. The overwhelming majority of studies reported that the interventions were at least partially effective, and two were shown to be cost-effective (Aas et al., 2019; Shearer et al., 2018). A cost-effectiveness analysis by Howarth et al (2016) suggested that for behavioural outcomes, a psychoeducational intervention delivered to parent and child in parallel is likely to be cost-effective among the interventions that they compared if willingness to pay was approximately £8000 (ICER = 3722 per SMD.

No evidence of economic effectiveness of traditional treatments, therapy or interventions was found for the period of the COVID-19 pandemic (2020-2022), when access to traditional group therapy was limited as the groups were either reduced in size, cancelled or moved online in response to the COVID-19 pandemic (Arnold and Burlingame, 2021). There is a real danger that a cohort of children/young people who have been affected by witnessing DVA during the COVID-19 pandemic have not been identified in the usual manner (e.g., by school staff or social workers) and therefore not been treated to prevent mental health issues in the future. Future publications may shed further light on this issue.

### 3.2 Strengths and limitations of the available evidence

The studies included in this review had several limitations related to internal and external validity. The main limitation was the concern about generalisability with small sample sizes. There is limited capacity for research to evaluate the effectiveness of interventions and support as there is a complex jigsaw of factors that influence the short-term and long-term consequences of experiencing DVA. For example, limited details about the possibility that the same participant both witnessed and experienced abuse was not seriously considered in the studies.

There was a paucity of studies including the outcomes of educational attainment, school/college attendance and school/college functioning. It was also unknown how many of the interventions described were co-produced by the children and young people that they aimed to support but there is a growing body of evidence that children’s voices should be heard and they are not passive bystanders but active sentient social actors in the unfolding story of their ongoing development (Øverlien and Holt, 2019).

### 3.3 Implications for policy and practice

#### Policy Implications

- Economic evaluations have found preliminary evidence that cognitive therapy is a cost-effective intervention to treat children and adolescents with PTSD.
- Future interventions should be co-produced with relevant stakeholders and patient and public members (including children and young people).

### 3.4 Strengths and limitations of this Rapid Review

#### 3.4.1 Strengths

The strength of this RR lies in that it identified relevant controlled trials and guidance documents. The RR provides a timely update on the evidence presented previously by Howarth and colleagues (Howarth et al., 2016). The evidence provided by Howarth and colleagues only reported on costs and did not identify any literature on the cost-effectiveness of interventions in this area. Our rapid review builds on this evidence by presenting the findings of two full economic evaluations that reported favourable cost-effectiveness findings for cognitive therapy interventions as well as identifying other clinical effectiveness studies of interest.

#### 3.4.2 Limitations

The main limitations of the included studies were that, in many of them the sample sizes were small, which impacts the statistical power of the study and, consequently, the reliability of the study findings. Some studies also had no non-treatment control groups making it difficult to know if the change would have occurred without intervention (Overbeek et al., 2013; Pernebo et al., 2019, 2018).

The review was limited by the number of published economic evaluations in this area. Consequently, we were unable to provide definitive conclusions regarding the cost-effectiveness of interventions for children and young people who have witnessed domestic abuse. There is a need for more robust economic evaluations, such as the cost-effectiveness of all types of interventions that may improve the outcomes for children and young people who have been exposed to DVA. Also, the longer-term impacts of interventions to reduce the harmful effects of witnessing DVA should be estimated in future research, given the life course impact.

It was not possible to provide conclusive appraisal of the costing studies included in this rapid review as there is no standard critical appraisal checklist/tool for these studies at present (Xu et al., 2021). In the absence of a standardised checklist for costing studies, the review team appraised the costing studies using the CHEERS checklist in line with other health economists (Xu et al., 2021). However, the nature of the questions were not always appropriate for the costing studies (Husereau et al., 2022).

## Data Availability

All data produced in the present study are available upon reasonable request to the authors

## Abbreviations

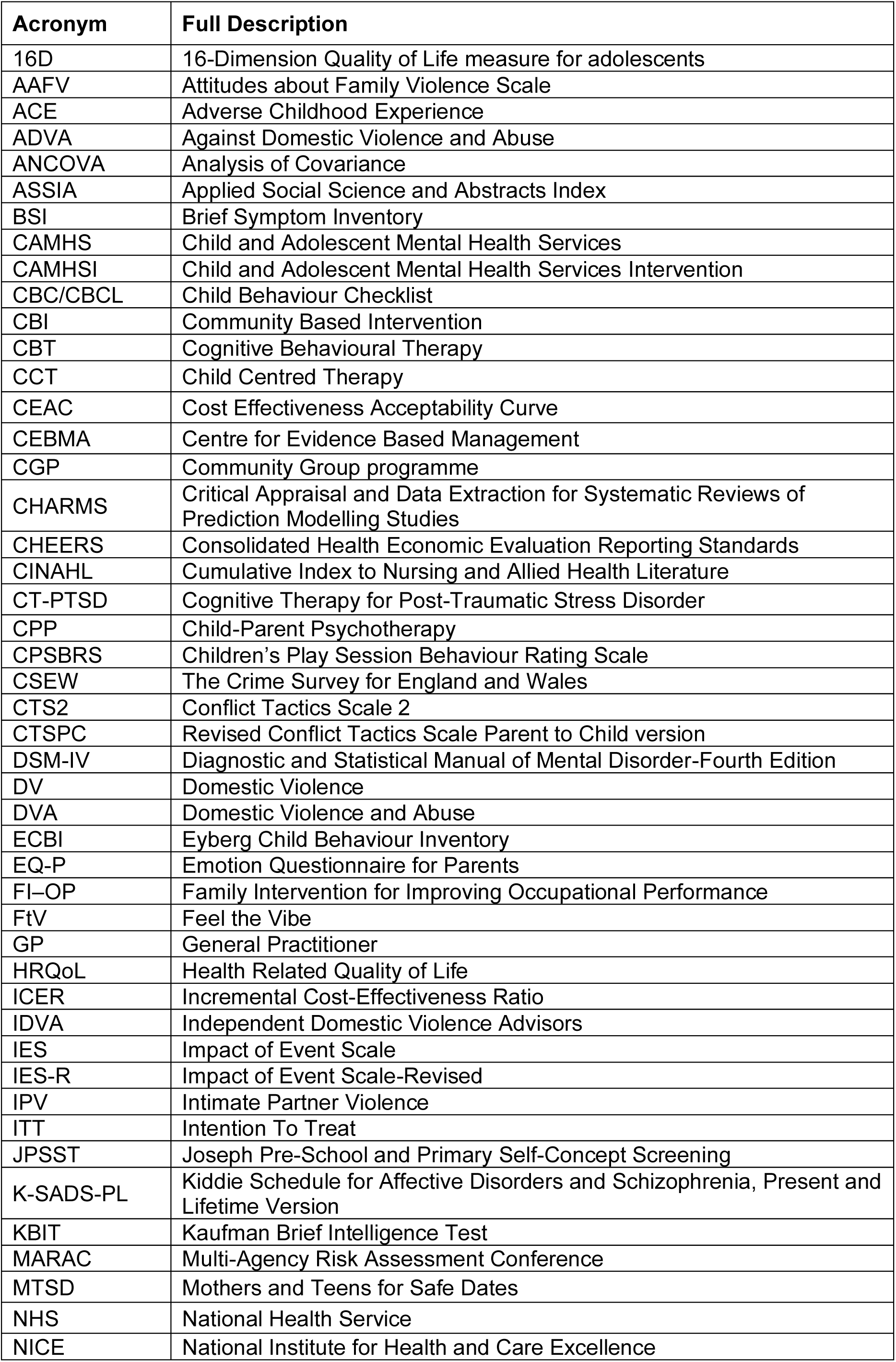
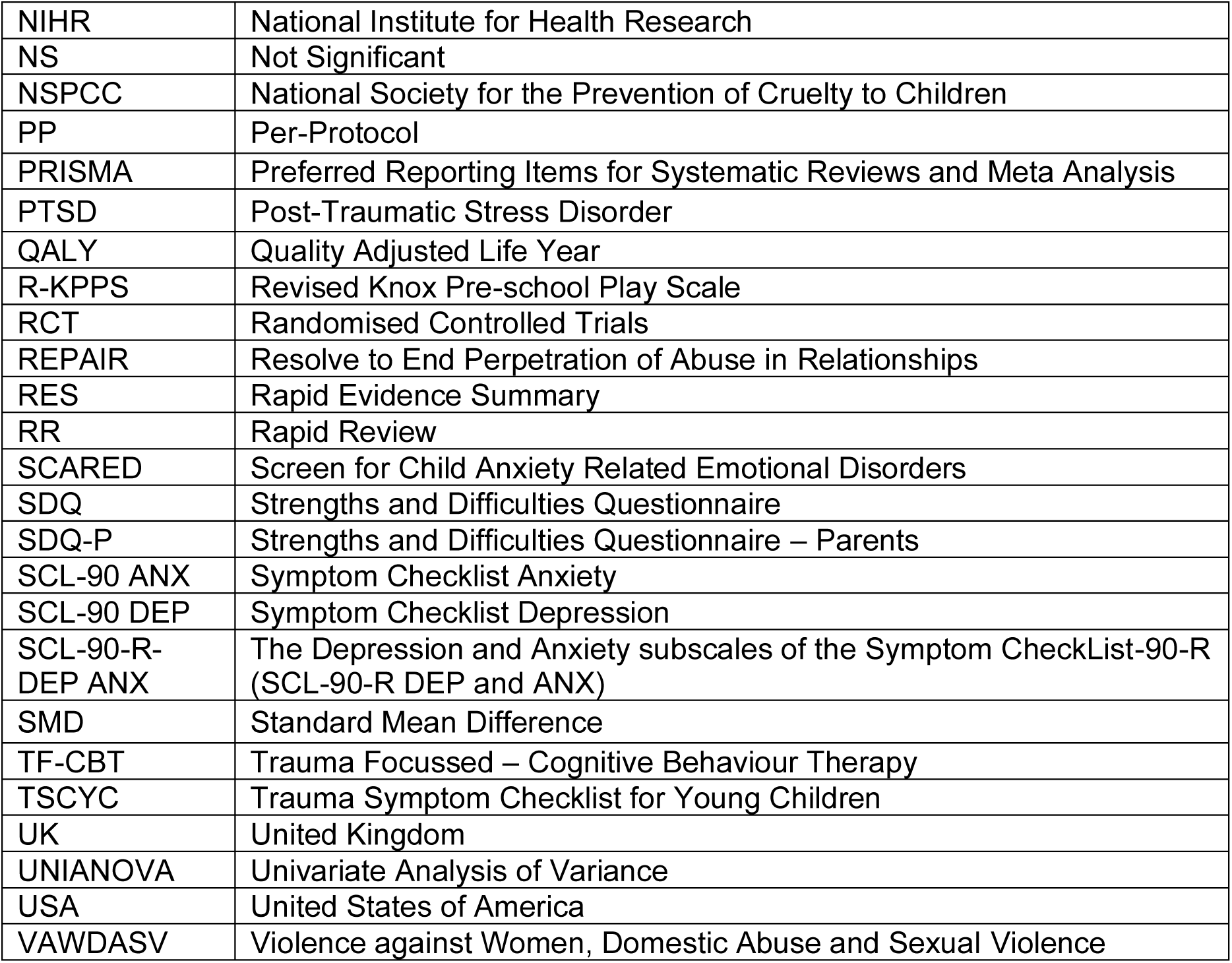

Abbreviations:
| Acronym | Full Description |
| --- | --- |
| 16D | 16-Dimension Quality of Life measure for adolescents |
| AAFV | Attitudes about Family Violence Scale |
| ACE | Adverse Childhood Experience |
| ADVA | Against Domestic Violence and Abuse |
| ANCOVA | Analysis of Covariance |
| ASSIA | Applied Social Science and Abstracts Index |
| BSI | Brief Symptom Inventory |
| CAMHS | Child and Adolescent Mental Health Services |
| CAMHSI | Child and Adolescent Mental Health Services Intervention |
| CBC/CBCL | Child Behaviour Checklist |
| CBI | Community Based Intervention |
| CBT | Cognitive Behavioural Therapy |
| CCT | Child Centred Therapy |
| CEAC | Cost Effectiveness Acceptability Curve |
| CEBMA | Centre for Evidence Based Management |
| CGP | Community Group programme |
| CHARMS | Critical Appraisal and Data Extraction for Systematic Reviews of Prediction Modelling Studies |
| CHEERS | Consolidated Health Economic Evaluation Reporting Standards |
| CINAHL | Cumulative Index to Nursing and Allied Health Literature |
| CT-PTSD | Cognitive Therapy for Post-Traumatic Stress Disorder |
| CPP | Child-Parent Psychotherapy |
| CPSBRS | Children's Play Session Behaviour Rating Scale |
| CSEW | The Crime Survey for England and Wales |
| CTS2 | Conflict Tactics Scale 2 |
| CTSPC | Revised Conflict Tactics Scale Parent to Child version |
| DSM-IV | Diagnostic and Statistical Manual of Mental Disorder-Fourth Edition |
| DV | Domestic Violence |
| DVA | Domestic Violence and Abuse |
| ECBI | Eyberg Child Behaviour Inventory |
| EQ-P | Emotion Questionnaire for Parents |
| FI-OP | Family Intervention for Improving Occupational Performance |
| FtV | Feel the Vibe |
| GP | General Practitioner |
| HRQoL | Health Related Quality of Life |
| ICER | Incremental Cost-Effectiveness Ratio |
| IDVA | Independent Domestic Violence Advisors |
| IES | Impact of Event Scale |
| IES-R | Impact of Event Scale-Revised |
| IPV | Intimate Partner Violence |
| ITT | Intention To Treat |
| JPSST | Joseph Pre-School and Primary Self-Concept Screening |
| K-SADS-PL | Kiddie Schedule for Affective Disorders and Schizophrenia, Present and Lifetime Version |
| KBIT | Kaufman Brief Intelligence Test |
| MARAC | Multi-Agency Risk Assessment Conference |
| MTSD | Mothers and Teens for Safe Dates |
| NHS | National Health Service |
| NICE | National Institute for Health and Care Excellence |
| NIHR | National Institute for Health Research |
| NS | Not Significant |
| NSPCC | National Society for the Prevention of Cruelty to Children |
| PP | Per-Protocol |
| PRISMA | Preferred Reporting Items for Systematic Reviews and Meta Analysis |
| PTSD | Post-Traumatic Stress Disorder |
| QALY | Quality Adjusted Life Year |
| R-KPPS | Revised Knox Pre-school Play Scale |
| RCT | Randomised Controlled Trials |
| REPAIR | Resolve to End Perpetration of Abuse in Relationships |
| RES | Rapid Evidence Summary |
| RR | Rapid Review |
| SCARED | Screen for Child Anxiety Related Emotional Disorders |
| SDQ | Strengths and Difficulties Questionnaire |
| SDQ-P | Strengths and Difficulties Questionnaire – Parents |
| SCL-90 ANX | Symptom Checklist Anxiety |
| SCL-90 DEP | Symptom Checklist Depression |
| SCL-90-R-DEP ANX | The Depression and Anxiety subscales of the Symptom CheckList-90-R (SCL-90-R DEP and ANX) |
| SMD | Standard Mean Difference |
| TF-CBT | Trauma Focussed – Cognitive Behaviour Therapy |
| TSCYC | Trauma Symptom Checklist for Young Children |
| UK | United Kingdom |
| UNIANOVA | Univariate Analysis of Variance |
| USA | United States of America |
| VAWDASV | Violence against Women, Domestic Abuse and Sexual Violence |

## 5. RAPID REVIEW METHODS

**Table 5.1.** **Inclusion and exclusion criteria**

|  | **Inclusion criteria** | **Exclusion criteria** |
| --- | --- | --- |
| **Population** | Children and young people (up to age 18 years of age) who have been exposed to domestic violence or abuse | Children and young people not exposed to domestic violence or abuse and those who have experienced abuse themselves. |
| **Settings of the intervention** | Any settings. Examples may include:<br>Home<br>Shelter<br>School<br>Healthcare settings<br>Community | No setting will be excluded. |
| **Intervention** | Any interventions that aim to prevent or limit the adverse impact of children and young people exposed to domestic violence or abuse (interventions of any duration from any setting). Examples may include:<br>Advocacy<br>Counselling<br>Psychoeducation<br>Psychotherapy<br>Psychosocial<br>Play therapy<br>Parenting skills training | Any intervention for children and young people not related to domestic violence or abuse |
| **Counter intervention** | Any type of alternative intervention or usual care. | N/A |
| **Outcome measures** | **Child behaviour**<br>i. child behaviour disorders<br>ii. child behaviour symptoms<br>**Children's mental health**<br>i. depression<br>ii. anxiety<br>iii. self-harm<br>iv. PTSD<br>**School based outcomes**<br>i. School attainment<br>ii. School attendance<br>iii. School functioning<br>**Children's self-esteem, self-competence or self-efficacy**<br>**Quality of life**<br>**Social relationships** | Any outcome measures not related to domestic violence or abuse. |
|  | **Intervention of social services** (children taken into care, child protection services, care conferences, etc.).<br>**Cost-effectiveness of interventions** |  |
| **Study design** | RCTs and studies in which participants were allocated to receive an intervention, a control, or no intervention without randomisation. | All other study designs will be excluded, as well as letters, commentaries, and references with no abstract. |
| **Countries** | Worldwide | No countries will be excluded from the searches. |
| **Language of publication** | English | Languages other than English. |
| **Publication date** | 23 September 2015 onwards | Pre-23 September 2015 |
| **Publication type** | Published and preprint | Grey literature |
| **Other factors**<br>*Any other key points to note* | The rapid review will utilise the evidence from existing SRs and will be based primarily on an update of the Howarth et al (2016) Mixed Methods Systematic Review (Chapter 3). This SR included controlled clinical trials and economic evaluations to address the following sub-questions:<br>• What is the evidence that existing interventions are effective?<br>• What is the economic evidence that existing interventions are cost-effective? |  |

### 5.1 Eligibility criteria

The eligibility criteria including inclusion and exclusion criteria will be presented in Table 2.

### 5.2 Literature search

Key databases were searched for studies published between 23 September 2015 and January 2023 as the Howarth et al (2016) (Howarth et al., 2016) SR searched up to 23 September 2015. The date limitations will be applied to keep the search within the scope of a Rapid Review. The following search strategy was written for Medline via OVID and then adapted for CINAHL, EMBASE, PsychINFO, Cochrane, ASSIA:

Search strategy (Medline via OVID)

1. Child/
2. Adolescent/
3. (adolesc* or child* or boy* or girl* or infant* or juvenil* or teen* or young or youth*).tw
4. 1 OR 2 OR 3
5. Domestic violence/
6. Spouse abuse/
7. Intimate partner violence/
8. Battered women/
9. (abuse* adj3 (wom* or partner* or spous* or m*n or wife or wives or husband*)).tw
10. (violen* adj3 (wom* or partner* or spous* or m*n or wife or wives or husband*)).tw
11. (marital adj3 (violen* or abus*)).tw
12. (intimate adj3 partner adj3 (violen* or abus*)).tw
13. (violen* adj2 (home or hous* or household)).tw
14. (abus* adj2 (home or hous* or household)).tw
15. 5 OR 6 OR 7 OR 8 OR 9 OR 10 OR 11 OR 12 OR 13 OR 14
16. (expos* or exposure or witness*).tw
17. ((child* or children or adolesc*) adj3 (living with)).tw
18. 16 OR 17
19. 4 AND 14 AND 18
20. Costs and cost analysis/
21. Cost-Benefit Analysis/
22. (economic adj2 (evaluation or cost* or analysis)).tw
23. (cost adj2 (effectiveness or benefit or consequence* or utility* or manage*)).tw
24. 20 OR 21 OR 22 OR 23
25. 19 AND 24

HE terms suggested:

(Economic evaluation or cost economic or economic analysis or cost-effectiveness or cost-benefit or cost-consequence or cost-utility or cost benefit or economics or cost management).

Databases searched:

Medline

ASSIA

PsycINFO

CINAHL

Embase Cochrane Library

NIHR Centre for Reviews and Dissemination (CRD) database including NHS EED database

### 5.3 Study selection process

Using the Covidence data screening and data extraction software tool for systematic reviews, citations were screened on title and abstract by two members of the core BIHMR Rapid Review team. Full-text articles were then retrieved and further assessed for inclusion. Any queries regarding inclusion/exclusion were resolved by discussion between members of the review team.

### 5.4 Data extraction

The data was extracted from the included studies using a pre-defined data extraction tool developed to capture all relevant data. Extracted data included study details such as author, year, setting, aim, design, population, sample size, type of study, type of intervention, method of analysis, key findings, and author conclusions.

Included papers were distributed among six members of the BIHMR review team for data extraction. A sample of extracted studies was checked against the papers for accuracy by the review lead. A proportion of the papers (10%) were double extracted to check for discrepancies between reviewers.

### 5.7 Quality appraisal

The RCT studies were quality appraised using the JBI RCT checklist (The Joanna Briggs Institute, 2020), and economic analysis papers, including the cost data studies, were quality appraised using the CHEERS Checklist (Husereau et al., 2022). Each study was quality appraised by a member of the BIHMR team, and the findings were then double assessed by another member of the BIHMR team. The quality appraisal tables can be found in Section 6 of this rapid review report.

### 5.8 Synthesis

Study characteristics and results are presented in tables 6.2.1-6.2.6, and the findings of this RR are presented narratively (Mishler, 1995). Identified key themes were used to structure the summary, and types of interventions were grouped according to intervention type, and then sub-grouped into intervention type and main outcome of interest.

### 5.9 Assessment of body of evidence

The overall body of evidence was described narratively, assessing specific aspects of the included studies, such as imprecision, inconsistency, indirectness, and publication bias. These assessments were limited to the primary outcomes of effectiveness and cost-effectiveness.

## 6. EVIDENCE

### 6.1 Study selection flow chart

**Figure 6.1:**
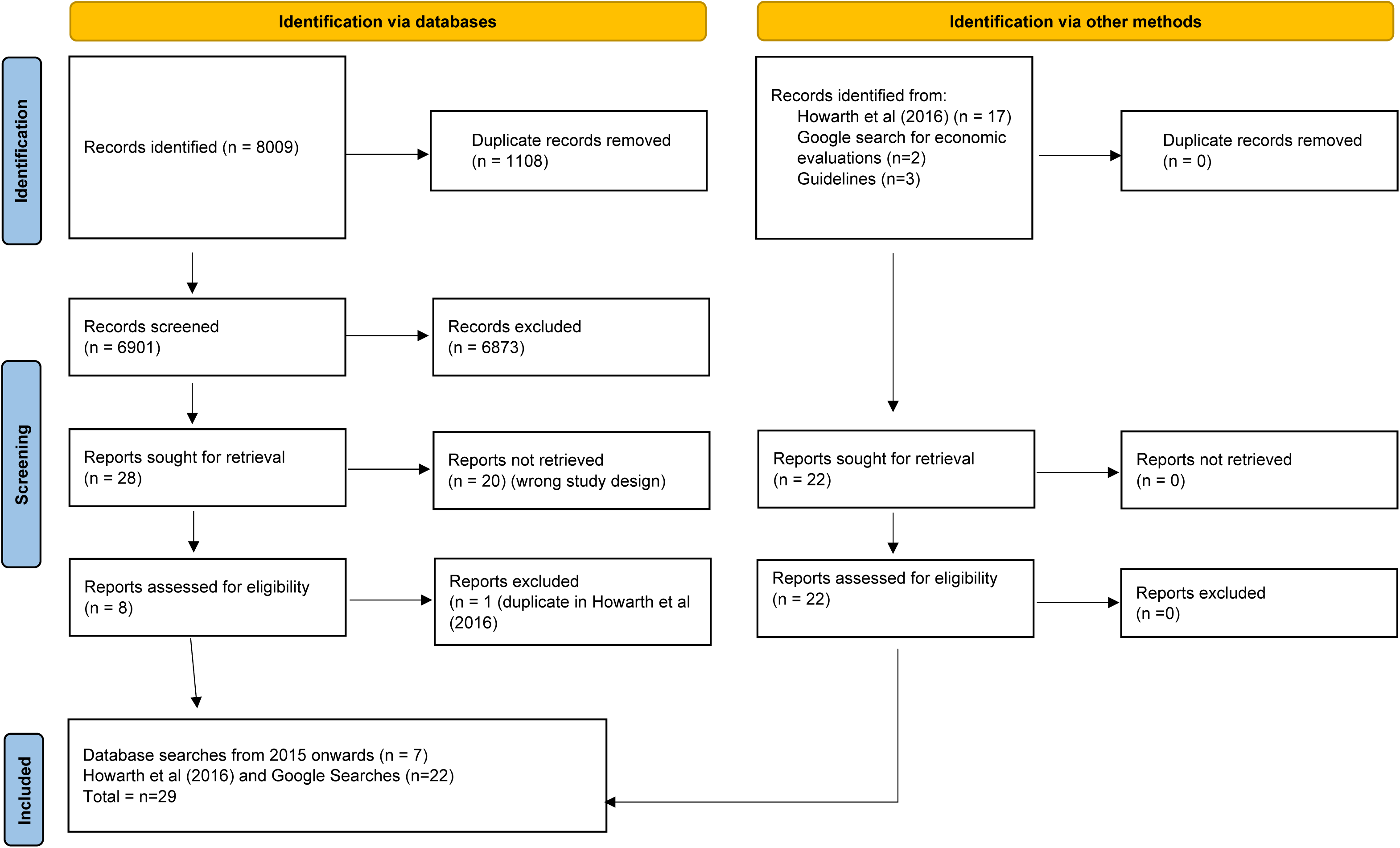
PRISMA 2020 flow diagram of included studies (Page et al., 2021)

### 6.2 Data extraction tables

Data extraction tables are shown below (see Tables 6.2.1 – 6.2.6).

**Table 6.2.1.**
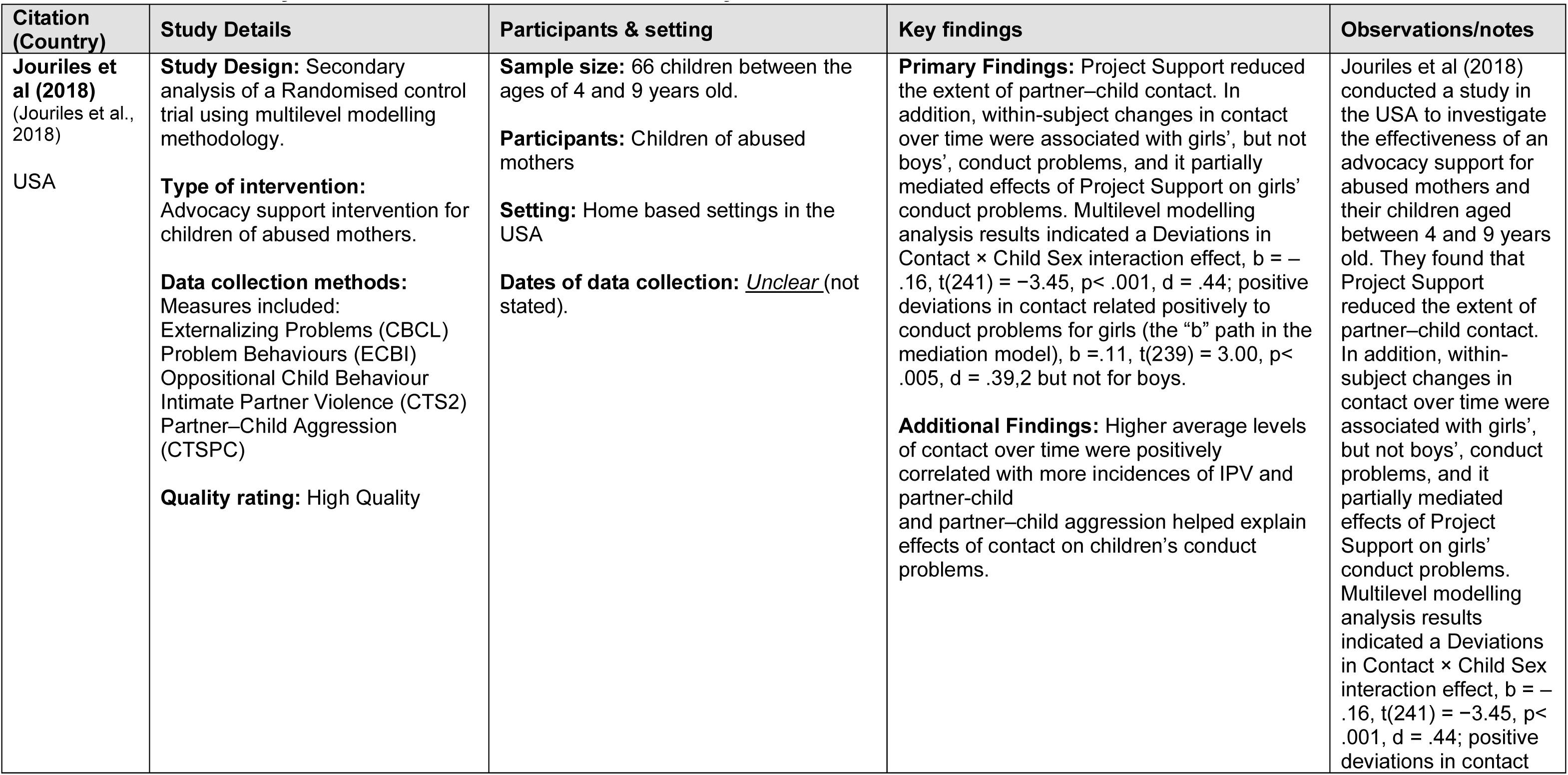
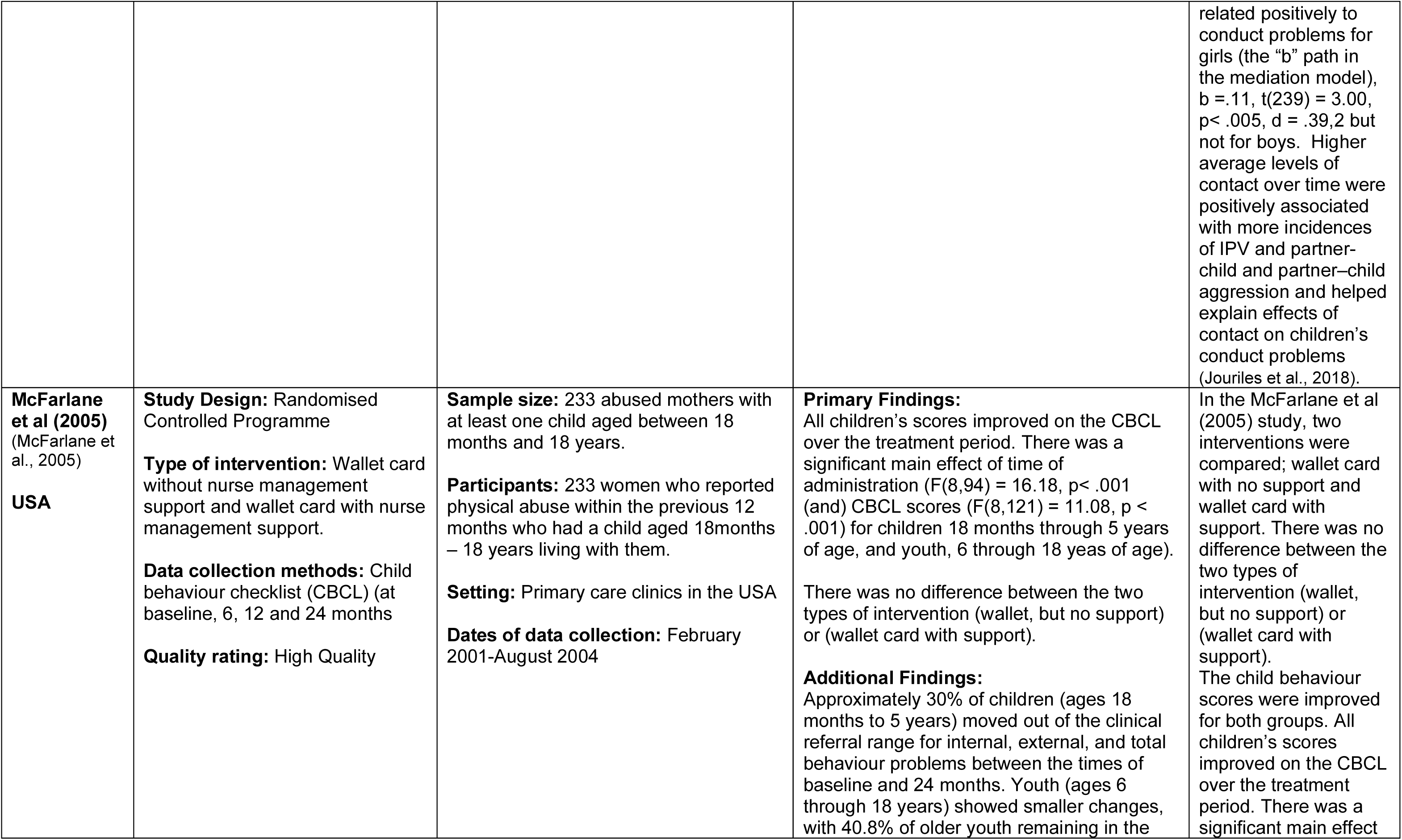
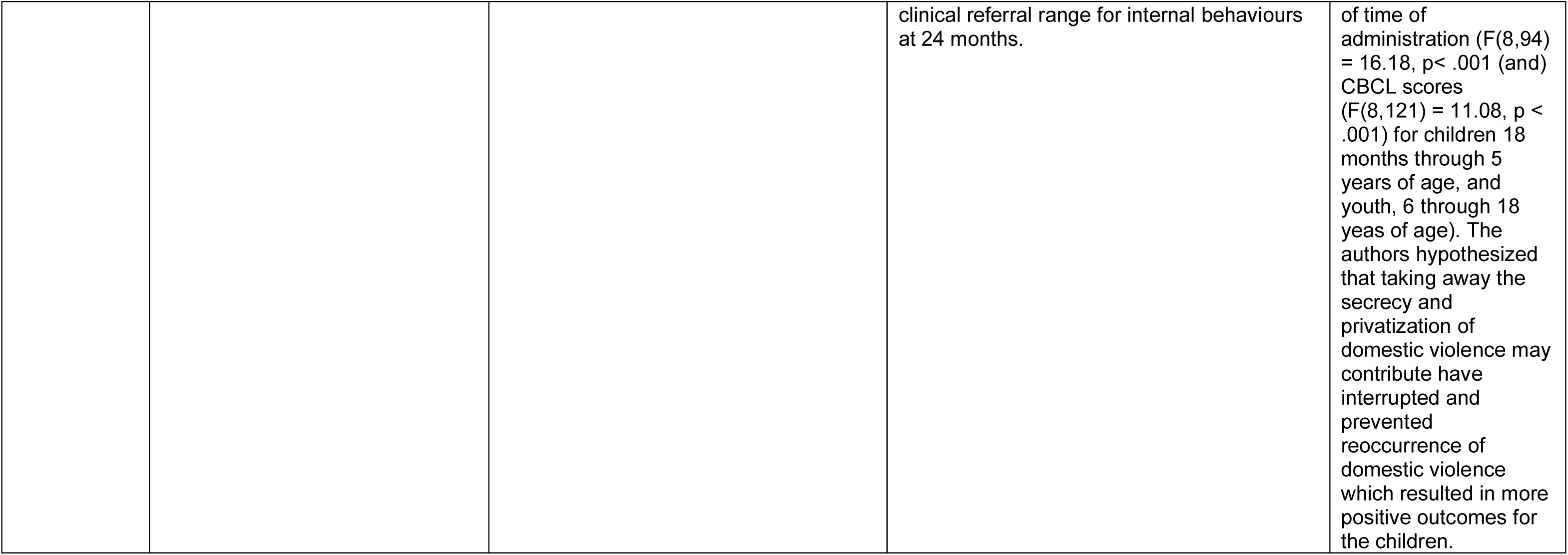
Summary of clinical effectiveness of Advocacy Services

**Table 6.2.2:** Summary of clinical effectiveness of Psychoeducation

| Citation (Country) | Study Details | Participants & setting | Key findings | Observations/notes |
| --- | --- | --- | --- | --- |
| **Kot et al (1998)**<br>(Kot et al., 1998)<br>**Canada** | **Study Design:** Randomised controlled study<br>**Type of intervention:**<br>Intervention – Child centred play therapy sessions in a period of 12 days to three weeks.<br>Control: Usual care group | **Sample size:** 22 children (6 girls and 5 boys). Mean age 6.9 years.<br>**Participants:** Children between the ages of 4 and 10 years old. 11 children in the intervention group and 11 children in the control group.<br>**Setting:** Community setting in Canada | **Primary Findings:**<br>ANCOVA analyses were conducted, and it was found that children in the experimental group scored significantly higher than the children in the control group on self-concept as measured by the JPPSST = Joseph Pre-School and Primary Self-Concept Screening Test: $F(1, 19) = 48.96, p < .001$. Children in the experimental group scored significantly higher than children in the control group in the CPSBRS areas of Physical Proximity, F | Kot et al (1998) (Kot et al., 1998) investigated the child centred play therapy with child witnesses of domestic violence, and found that play therapy within a period of three weeks was effective and behaviours and self-concepts |
|  | **Data collection methods:**<br>Child's behaviour. The Child Behaviour Checklist (CBCL) (Achenbach, 1991) consists of 118 items designed to record in a standardized format the behaviours and competencies of children as reported by their parents. This study used the following scales: Internalizing Behaviour Problems, Externalizing Behaviour Problems, and Total Behaviour Problems.<br>Children's Play Session Behaviour Rating Scale (CPSBRS)<br>**Quality rating:** Moderate Quality | **Dates of data collection:** <u>Unclear</u> (data collected prior to publication in 1998) | (1, 19) = 13.561, $p < .01$; and Play Themes, $F(1, 19) = 12.18$, $p < .01$.<br>There was also a significant ($p < .05$) reduction in externalizing behaviour problems for the experimental group at post-test. The mothers of children in the experimental group perceived their children as less aggressive and as manifesting fewer delinquent behaviours such as lying, cheating, and swearing. This decrease in behaviour problems was perceived by the mothers to be particularly noteworthy.<br>**Additional Findings:**<br>One therapist described a four-year-old girl's progress in therapy: "Cindy first came into the playroom with both excitement and fears. She was excited by the wide range of toys available to her but was afraid of the spider, alligator, snake, and other wild animals. Cindy purposely stayed away from the scary animals. As several play sessions went by, Cindy re-enacted her scary escape from home after a violent episode, and she approached the scary animals again. She gradually moved from looking at them and touching them to picking the animals up. During the last two sessions, she picked up the scary animals and told them one by one, in an assured voice, "I am not afraid of you" and proceeded to throw them onto the floor". | improved. ANCOVA analyses were conducted, and it was found that children in the experimental group scored significantly higher than the children in the control group on self-concept as measured by the JPPSST = Joseph Pre-School and Primary Self-Concept Screening Test: $F(1, 19) = 48.96$, $p < .001$. Children in the experimental group scored significantly higher than children in the control group in the CPSBRS areas of Physical Proximity, $F(1, 19) = 13.561$, $p < .01$; and Play Themes, $F(1, 19) = 12.18$, $p < .01$.<br>There was also a significant ($p < .05$) reduction in externalizing behaviour problems for the experimental group at post-test. The mothers of children in the experimental group perceived their children as less aggressive and as<br>manifesting fewer delinquent behaviours such as lying, cheating, and swearing. This decrease in behaviour problems was perceived by the mothers to be particularly noteworthy. One therapist described a four-year-old girl's progress in therapy: "Cindy first came into the playroom with both excitement and fears. She was excited by the wide range of toys available to her but was afraid of the spider, alligator, snake, and other wild animals. Cindy purposely stayed away from the scary animals. As several play sessions went by, Cindy re-enacted her scary escape from home after a violent episode, and she approached the scary animals again. She gradually moved from looking at them and touching them to picking the animals up. During the last two<br>sessions, she picked up the scary animals and told them one by one, in an assured voice, "I am not afraid of you" and proceeded to throw them onto the floor". |
| **Graham-Bermann et al (2007)**<br>(Graham-Bermann et al., 2007)<br>**USA** | **Study Design:** Randomised Clinical Trial<br>**Type of intervention:** An intervention to reduce conduct problem among children of parents experiencing interpersonal violence<br>**Data collection methods:** Attitudes about Family Violence Scale (AAFV)<br>**Quality rating:** Moderate to Low Quality | **Sample size:** 66 families (mothers and children)<br>**Participants:** Children of mothers experiencing interpersonal violence (n=66 children between the ages between 4 to 9 years). Intervention group n=32 (project support) Comparison group n=34 (control)<br>**Setting:** Community setting in the USA<br>**Dates of data collection:** October 1996 to January 2000 | **Primary Findings:** During the intervention period, child conduct problems decreased in the Project Support group, $b = -.53$, $t(64) = -7.13$, $p < .001$, as well as in the comparison group, $b = -.30$, $t(64) = -5.16$, $p < .01$. However, they decreased more rapidly in the Project Support group than in the comparison group, $b = .23$, $t(64) = 2.78$, $p < .02$.<br>**Additional Findings:** Oppositional child behaviour decreased more slowly than other measures of child conduct problems, $b = .39$, $t(332) = 4.92$, $p < .001$. | Graham-Bermann et al (2007) conducted a randomised clinical trial to investigate the effectiveness of an intervention to reduce conduct problems among children of parents experiencing interpersonal violence in the USA.<br>During the intervention period, child conduct problems decreased in the Project Support group, $b = -.53$, $t(64) = -7.13$, $p < .001$, as well as in the comparison group, $b = -.30$, $t(64) = -5.16$, $p < .01$. However, they decreased more rapidly in the Project Support group than in the comparison group, $b = .23$, $t(64) = 2.78$, $p < .02$.<br>Oppositional child behaviour decreased more slowly than other<br>measures of child conduct problems, $b = .39$ , $t(332) = 492$ , $p < .001$ . |
| **Graham-Bermann et al (2015)**<br>(Graham-Bermann et al., 2015)<br>**USA** | **Study Design:** Randomised Controlled Trial (RCT)<br>**Type of intervention:** Psychoeducational intervention for children called the pre-kids club (PKC).<br>**Data collection methods:** Mothers were interviewed pre and post- intervention (approximately 5 weeks apart) and at 8-month follow-up<br>**Quality rating:** Moderate to High Quality | **Sample size:** 36 (intervention and 35 (control).<br>**Participants:** Children of abused mothers<br>**Setting:** Community settings in the USA<br>**Dates of data collection:** Took 2 years to conduct this study<br><u>Unclear</u> | **Primary Findings:**<br>The hypothesis that children in the intervention condition would show significant improvement in internalizing symptoms, relative to those in the no treatment comparison condition, was partially supported.<br>Effect sizes (Cohen's d) from Time 1 to Time 2 were .18 for the treatment group and .15 for the comparison group. Effect sizes of change from Time 2 to Time 3 were small (.01 for the treatment and -.07 for the comparison group).<br>**Additional Findings:**<br>ITT analyses indicated the program reduced internalizing problems for girls at follow- up. PP analyses indicated the program reduced internalizing problems for both boys and girls at post-intervention. In this study, child internalizing problems were significantly reduced through an intervention for the mother and the child. | **Graham-Bermann et al (2015)** (Graham-Bermann et al., 2015) **conducted an RCT in community settings in the USA with children of abused mothers**<br>The hypothesis that children in the intervention condition would show significant improvement in internalizing symptoms, relative to those in the no treatment comparison condition, was partially supported.<br>Effect sizes (Cohen's d) from Time 1 to Time 2 were .18 for the treatment group and .15 for the comparison group. Effect sizes of change from Time 2 to Time 3 were small (.01 for the treatment and -.07 for the comparison group). |
| **Overbeek et al (2013)**<br>(Overbeek et al., 2013)<br>**The Netherlands** | **Study Design:** Control groups study<br>**Type of intervention:** 9 group therapy sessions<br>**Data collection methods:** Post traumatic stress measures<br>**Quality rating:** High Quality | **Sample size:** 155 parents and children aged 6-12 years (55% boys)<br>**Participants:** Children aged between 6 years and 12 years old.<br>**Setting:** A community-based setting in The Netherlands<br>**Dates of data collection:** September 2009 – January 2012 | **Primary Findings:**<br>Child self-report: Depressive symptoms. Children's self-reports revealed a decrease in depressive symptoms over time from baseline to post-test, which was maintained to follow-up ($B = -6.72$, $t(1, 111.76) = -5.88$, $p < .001$), irrespective of condition. Both intervention and control groups scored similarly on 'structure' (adherence to the intervention/control) and 'positive attention' but the intervention group improved on 'Emotion differentiation and regulation', 'Emotion differentiation and regulation: IPV', 'Teaching general coping skills', 'Coping skills regarding IPV' and 'Sharing of experiences regarding IPV'.<br>**Additional Findings:**<br>Higher adherence was not associated with a larger difference in recovery between intervention and control conditions. | Overbeek et al (2013) investigated the post-traumatic stress of children aged 6 to 12 years in The Netherlands. They found that the 9-week group therapy intervention was effective in improving emotional and coping skills of children who had experienced post-traumatic stress as a consequence of IPV. Depressive symptoms were reduced ($p < .001$). |
| **Sullivan et al (2002)**<br>(Sullivan et al., 2002)<br>**USA** | **Study Design:** Randomised controlled study<br>**Type of intervention:** Strength based advocacy intervention for abused women and their children. The experimental intervention involved advocacy for mothers and their children and a 10-week support and education group for the children. Families in the experimental condition received the free services of a trained paraprofessional for 6 to 8 hours per week over 16 weeks. | **Sample size:** 78 families who had been users of a domestic violence shelter programme.<br>**Participants:** All participants were from a domestic violence shelter programme. All mothers had at least one child between 7 and 11 years old.<br>**Setting:** USA community setting<br>**Dates of data collection:** <u>Unclear</u> (before publication in 2002) | **Primary Findings:**<br>Children in the experimental condition reported significantly higher self-competence in several domains (global self-worth, physical appearance and athletic sub-scale) compared to children in the control group ($p < .05$).<br>**Additional Findings:**<br>The authors noted that the measures used to measure intervention effectiveness were not sensitive enough. | The RCT by Sullivan et al (2002) conducted in the USA found that the strength-based intervention for abused women and their children was moderately effective. Children in the experimental condition reported significantly higher self-competence in several domains (global self-worth, physical appearance and athletic sub-scale) compared to children |
|  | **Data collection methods:** Qualitative interviews and Measures including: Measures of abuse (developed by the authors).<br>**Quality rating:** Moderate Quality |  |  | in the control group ($p < .05$). The authors noted that the measures used to measure intervention effectiveness were not sensitive enough. |
| **Wagar and Rodway (1995)**<br>(Wagar and Rodway, 1995)<br>**Canada** | **Study Design:** Randomised controlled study<br>**Type of intervention:** Group treatment including education and knowledge about how to keep safe (timeframe of the intervention was not mentioned in the text, and his therefore unknown).<br>**Data collection methods:** Child Witness to Violence Questionnaire<br>Parent to child questionnaire (to measure type of abuse)<br>Parent interview questionnaire (to gain socio demographic information on the children's families)<br>Self-report evaluations at the end of the programme<br>**Quality rating:** Moderate to Low Quality | **Sample size:** 42 children from Canada<br>**Participants:** Of the 52 children referred by several community service agencies, 42 were eligible for the groups. There were 22 children in the 8- to 10- year-old-age bracket and 20 children in the 11- to 13-year-old age bracket.<br>**Setting:** Canadian healthcare setting<br>**Dates of data collection:** <u>Unclear</u> (pre-publication in 1995) | **Primary Findings:** The analysis of co-variance was used to test the hypothesis that children in the treatment group would be different on the three dependent variables: (1) attitudes and responses to anger, (2) knowledge of safety and support, (3) sense of responsibility for parents and for the violence, than the children in the control group. The results of the analysis indicated that the children in the treatment group when compared to children in the control group showed significantly higher post-test scores on two of the dependent variables after removing the effects of the pre-test scores: attitudes and responses to anger ($F$ Main effect=8.23, $p < .01$, and sense of responsibility for parents and the violence ($F$ Main effect=4.50, $p < .05$).<br>Knowledge of safety and support did not differ between the treatment and control groups ($p > .05$).<br>**Additional Findings:**<br>In self-evaluative reports, it was found that children would demonstrate destructive behaviour. It was also found in this study that children who tended to be less aggressive in peer interaction, according to report cards<br>and parents, appeared to exhibit self-harming behaviours, such as taking high risks in potentially dangerous situations. Other children were more aggressive with their peers. May children noted how this provided “a relief in the tension”. | Wagar and Rodaway (1995) conducted a randomised controlled trial in Canada with a group treatment including education and knowledge about how to keep safe, and a control condition. They found that children in the treatment condition had improved scores on attitudes and attitudes and responses to anger ($F$ Main effect=8.23, $p < .01$, and sense of responsibility for parents and the violence ($F$ Main effect=4.50, $p < .05$). |
| **Foshee et al (2015)**<br>**USA**<br>(Foshee et al., 2015)<br>**USA** | **Study Design:** RCT<br>**Type of intervention:**<br>Dating training for adolescents of abused mothers (Mothers and Teens for Safe Dates, MTSD) Families in the treatment group were mailed a program booklet every 2 weeks.<br>Families in the control group were not sent any program materials.<br>**Data collection methods:**<br>Dating abuse outcomes were measured including cyber dating abuse, sexual dating abuse and physical dating abuse.<br>**Quality rating:** High Quality | **Sample size:** 409 adolescents (mother and adolescent pairs)<br>**Participants:** Adolescents (teenage children of abused mothers aged between 12 and 16 years). 64% were female.<br>**Setting:** Community settings in the USA<br>**Dates of data collection:**<br>Previous to acceptance of the paper for publication in 2014. | **Primary Findings:** The MTSD program had significant program effects on the perpetration of cyber abuse ($p < .05$), but not for adolescents who had average or low exposure to DVA ($p > 0.1$). There were no program effects on the perpetration of physical or sexual dating abuse.<br>**Additional Findings:**<br>The amount of exposure the adolescent had to domestic violence was also not associated with the number of intervention booklets completed. | Foshee et al (2015) conducted an RCT in the USA to investigate the effectiveness of the MTSD program had significant program effects on the perpetration of cyber abuse ($p < .05$), but not for adolescents who had average or low exposure to DVA ($p > 0.1$) There were no program effects on the perpetration of physical or sexual dating abuse. |
| **Sargent et al (2016)**<br>(Sargent et al., 2016)<br>**USA** | **Study Design:** RCT<br>**Type of intervention:**<br>Intervention: An on-line programme called Change A Life<br>(This interactive, web-based program uses short video clips, quizzes, and informational drop-downs to inform users about the | **Sample size:** Community participants n=110<br>University students n=146<br>**Participants:** Participants were recruited from two demographically different samples: a community sample from a large metropolitan area and a college student sample from a mid-size, private university. | **Primary Findings:** This RCT investigated the effects of an online educational program in increasing knowledge about children's exposure to domestic violence (DVA). Both community group and undergraduate intervention group improved their knowledge of DVA as well as their self-efficacy for helping children who have experienced DVA after being involved in the Change A Life intervention ($P < .001$). The control group | Sargent et al (2016) conducted an RTC in the USA to investigate the effects of an online educational program in increasing knowledge about children's exposure to domestic violence (DVA). Both community group and |
|  | prevalence and impact of children's exposure to DVA, as well as step-by-step training on how to help children exposed to DVA to foster resilience).<br>**Control group:** Participants assigned to the control arm completed an online program offered by the Alzheimer's Association titled 'Know the 10 Signs: Early Detection Matters'. It includes information about disease detection, recognizing symptoms, and what to do to help affected individuals, presented in a 35-min video.<br>**Data collection methods:**<br>Knowledge About<br>Consequences and how to Help<br>Children Exposed to DVA<br>Self-Efficacy to Help Children<br>Exposed to DVA<br>History of Childhood<br>DVAExposure<br>**Quality rating:** High Quality | **Setting:** Southwest region of the USA<br>**Dates of data collection:** Pre-January 2016. | completed an on-line programme from the Alzheimer's Association did not show an improvement in their knowledge of DVA ($P > 0.1$).<br>**Additional Findings:** Neither participant gender nor prior exposure to domestic violence moderated the intervention effects. | undergraduate intervention group improved their knowledge of DVA as well as their self-efficacy for helping children who have experienced DVA after being involved in the Change A Life intervention ($P < .001$). The control group completed an on-line programme from the Alzheimer's Association did not show an improvement in their knowledge of DVA ($P > 0.1$). Neither participant gender nor prior exposure to domestic violence moderated the intervention effects (Sargent et al., 2016) |
| **Van Rosmalen-Nooijens et al (2017)**<br>(Van Rosmalen- | **Study Design:** RCT<br>**Type of intervention:**<br>**Intervention:** Internet-based self-support for Adolescents and young adults exposed to family violence (FtV intervention)<br>**Control:** minimally enhanced usual care | **Sample size:** 17 participants (n=8 intervention arm and n=9 control arm).<br>**Participants:** Adolescents and young adults aged 12-25 years old<br>**Setting:** Home based setting in The Netherlands | **Primary Findings:**<br>Seventeen participants (intervention: n=8, control: n=9) completed all questionnaires. Mixed model analysis showed significant differences between groups on the SCL-90 DEP ($P=.04$) and ANX ($P=.049$) subscales between 6 and 12 weeks after participation started. UNIANOVA showed no significant differences. Pre-post paired sample t-tests | Van Rosmalen-Nooijens et al (2017) conducted an RCT in the Netherlands and found that the Seventeen participants (intervention: n=8, control: n=9) completed all |
| Nooijens et al., 2017)<br>**The Netherlands** | **Data collection methods:**<br>Impact of Event Scale (IES)<br>Symptom Checklist Depression (SCL-90 DEP)<br>Symptom Checklist Anxiety SCL-90 ANX)<br>**Quality rating:** High Quality | **Dates of data collection:** June 2012 to July 2014. | showed significant improvements after 12 weeks for the SCL-90 DEP ($P=.03$) and ANX ($P=.046$) subscales.<br>On the Impact of Event Scale (IES), there was no significant difference between the intervention group and the control group at the final follow-up, $P > 0.1$.<br>**Additional Findings:**<br>All participants involved with the internet-based intervention felt safe. | questionnaires. Mixed model analysis showed significant differences between groups on the SCL-90 DEP ($P=.04$) and ANX ($P=.049$) subscales between 6 and 12 weeks after participation started. UNIANOVA showed no significant differences. Pre-post paired sample t-tests showed significant improvements after 12 weeks for the SCL-90 DEP ($P < .03$) and ANX ($P < .05$) subscales.<br>On the Impact of Event Scale (IES), there was no significant difference between the intervention group and the control group at the final follow-up, $P > 0.1$. In terms of additional findings, all participants involved with the internet-based intervention felt safe. (Van Rosmalen-Nooijens et al., 2017). |

**Table 6.2.3:** Summary of clinical effectiveness of Cognitive Behavioural Therapy and/or Psychotherapy

| Citation (Country) | Study Details | Participants & setting | Key findings | Observations/notes |
| --- | --- | --- | --- | --- |
| **Cohen et al (2011)**<br>(Cohen et al., 2011)<br>**USA** | **Study Design:** Randomised controlled trial conducted using blinded evaluators.<br>**Type of intervention:** 8 sessions of Trauma Focussed Cognitive behaviour therapy (CBT) versus usual care (child-centred therapy, CCT)<br>**Data collection methods:** The PTSD symptom clusters were assessed using the Kiddie Schedule for Affective Disorders and Schizophrenia, Present and Lifetime Version (K-SADS-PL). Children's anxiety symptoms were assessed using the Screen for Child Anxiety Related Emotional Disorders (SCARED), a self-report measure of non-PTSD anxiety. Children's depressive symptoms were assessed using the Children's Depression Inventory. Children's total behaviour problems were assessed using the Child Behaviour Checklist, a parent report instrument. Cognitive functioning was assessed using the Kaufman Brief Intelligence Test, a brief measure of verbal and nonverbal intelligence. | **Sample size:** (n=124) 7–14-year-old children<br>**Participants:** 7–14-year-old children (n=124)<br>N=61 male and n=63 female participants. N=64 received TF-CBT, n=60 received CCT.<br>**Setting:** Community setting in the USA<br>**Dates of data collection:** Between September 2005 and June 2009 | **Primary Findings:** Children completing TF-CBT had significantly greater improvement than children completing CCT in K-SADS-PL total score (1.67; –0.08 to 3.4) and Reaction Index score (–7.58; –0.79 to –14.38) and in K-SADS-PL hyperarousal score (–0.81; –0.03 to –1.59) and anxiety score (–7.36; –1.06 to –13.67).<br>**Additional Findings:** Following the intervention, children could feel safer in the face of ongoing danger; for example, differentiating between real and generalized fears, learning safety and relaxation strategies, and talking directly to the mother about IPV experiences. These interventions may have been particularly effective for hyperarousal symptoms, such as generalized vigilance, irritability, difficulty sleeping, and anger, and for decreasing avoidance about talking about IPV experiences with the mother. | Cohen et al (2011) conducted an RCT in community settings in the USA to investigate the effectiveness of an 8-week CBT intervention for n=124 7- to 14-year-olds. They found that the children in the intervention group made greater improvements than children in the usual care groups. Children completing TF-CBT had significantly greater improvement than did children completing CCT in K-SADS-PL total score (1.67; –0.08 to 3.4) and Reaction Index score (–7.58; –0.79 to –14.38) and in K-SADS-PL hyperarousal score (–0.81; –0.03 to –1.59) and anxiety score (–7.36; –1.06 to –13.67). Following the intervention, children could feel safer in the face of ongoing |
|  | **Quality rating:** High Quality |  |  | danger; for example, differentiating between real and generalized fears, learning safety and relaxation strategies, and talking directly to the mother about IPV experiences. |
| **Lieberman et al (2005)**<br>(Lieberman et al., 2005)<br>**USA** | **Study Design:** Randomised controlled trial<br>**Type of intervention:** Child-Parent Psychotherapy (CPP) and usual care over 50 weeks.<br>**Data collection methods:** Child Behaviour Checklist (CBCL)<br>**Quality rating:** High Quality | **Sample size:** 75 children aged between 3 and 5 years old<br>**Participants:** Children between 3 and 5 years old (39 boys and 36 girls)<br>**Setting:** Community setting in USA<br>**Dates of data collection:** *Unclear* (not reported in the publication). | **Primary Findings:** Analyses of CBCL Total scores showed a significant group 3-time interaction, $F_{1,61} = 5.77$, $p < .05$, $d = 0.24$, with follow-up analyses revealing that only the CPP group evidenced significant intake-post-test reductions: $t(34) = 2.86$, $p < .01$. To examine whether error was introduced because some children completed the CBCL 2-3 at intake and the CBCL 4-18 at post-test, analyses were repeated with only the children who completed the CBCL 4-18 at intake and post-test. These analyses also resulted in a significant interaction effect ($F_{1,31} = 4.72$, $p < .05$, $d = 0.64$), with follow-up analyses confirming that only the CPP group showed significant reductions in behaviour problems (CPP: intake mean = 60.32, SD = 9.00; post-test mean = 54.16, SD = 8.71, $t(18) = 3.10$, $p < .01$; comparison: intake mean = 58.86, SD = 8.82; post-test mean = 59.64, SD = 13.11).<br>**Additional Findings:**<br>The overall findings highlight the importance of including the mother as an integral partner in the treatment of pre-schoolers' traumatic stress symptoms. | Lieberman et al (2005) (Lieberman et al., 2005) investigated the effectiveness of child-parent psychotherapy versus usual care over 50 weeks in community settings in the USA. Analyses of CBCL Total scores showed a significant group 3-time interaction, $F_{1,61} = 5.77$ , $p < .05$ , $d = 0.24$ , with follow-up analyses revealing that only the CPP group evidenced significant intake-post-test reductions: $t(34) = 2.86$ , $p < .01$ . To examine whether error was introduced because some children completed the CBCL 2-3 at intake and the CBCL 4-18 at post-test, analyses were repeated with only the children who completed the CBCL 4-18 at intake and post-test. These analyses also resulted in a significant interaction effect ( $F_{1,31} = 4.72$ , $p < .05$ , $d = 0.64$ ), with follow-up analyses confirming that only the CPP group showed significant reductions in behaviour problems (CPP: intake mean = 60.32, SD = 9.00; post-test mean = 54.16, SD = 8.71, $t(18) = 3.10$ , $p < .01$ ; comparison: intake mean = 58.86, SD = 8.82; post-test mean = 59.64, SD = 13.11). |
| **McWhirter et al (2011)**<br>(McWhirter, 2011)<br>**USA** | **Study Design:** Randomised Controlled Study<br>**Type of intervention:**<br>Intervention 1: emotion focussed orientated group therapy<br>Intervention 2: goal focussed oriented group therapy.<br>**Data collection methods:**<br>**Emotional barometer** – with a seven-point Likert scale.<br>**Psychosocial measures** – 1) peer conflict, 2) family conflict, 3) self-esteem were all measured | **Sample size:** 46 women and their children ($n=48$) aged between 6 and 12 years old.<br>**Participants:** Temporary homeless families (mothers and their children).<br>**Setting:** Community settings in the USA<br>**Dates of data collection:** <u>Unclear</u> (pre-publication in 2011). | **Primary Findings:**<br>A series of 2 (treatment) x 2 (time) repeated measures analyses revealed main effects for state of emotional well-being, $F(1, 46) = 7.00$, $p < .05$, $\eta^2 = .13$; peer conflict, $F(1, 46) = 4.97$, $p < .05$, $\eta^2 = .16$; family conflict, $F(1, 46) = 22.27$, $p < .05$, $\eta^2 = .43$; and self-esteem, $F(1, 46) = 7.87$, $p < .05$, $\eta^2 = .24$. Children in both groups reported decreases in family and peer conflict and increases in state of emotional well-being and self-esteem.<br>**Additional Findings:** | In the randomised controlled study by McWhirter et al (2011) temporary homeless families in the USA (mothers and their children) were asked to complete emotional and psychosocial measures. The child participants fundamentally responded in positive ways to both interventions (emotion |
|  | with a single item (five-point Likert scale) designed for this study.<br>**Quality rating:** High Quality |  | The multiplicity of concerns and symptoms experienced by women and children subsequent to IPV exposure demands further development and research involving effective practice-based community interventions designed to better meet the needs of homeless women and children. | and goal focussed group therapy). The results of this study are congruent with studies that demonstrate the efficacy of family-based interventions involving the child with his or her mother following intimate partner violence. Multicomponent approaches involving mothers together with children are successful in improving attitudes about violence and reducing aggression among children exposed to domestic violence and abuse. |
| **Pernebo et al (2018)**<br>(Pernebo et al., 2018)<br>**Sweden** | **Study Design:** The study used a quasi-experimental design with assessment before (T1) and after termination of (T2) the intervention.<br>**Type of intervention:** Psychotherapy and psychoeducational interventions<br>**Data collection methods:** Conflict Tactics Scale (CTS2) Strengths and Difficulties Questionnaire (SDQ-P) Trauma Symptom Checklist for Young Children (TSCYC) Emotion Questionnaire for parents (EQ-P) | **Sample size:** n=31 in the psychoeducation group and n=19 CAMHS psychotherapy group<br>**Participants:** Children exposed to intimate partner violence<br>**Setting:** Community based intervention in Sweden<br>**Dates of data collection:** 2013–2015 | **Primary Findings:** The mothers in the Community Based Intervention (CBI) reported a significant reduction in their child's emotional symptoms (SDQ-P; d=0.34), in total post-traumatic stress (TSCYC; d=0.35), and in intrusive symptoms (TSCYC; d=0.40). Mothers in the CBI additionally reported a significant decrease in impact scores (SDQ-P; d=0.62). The mothers in the CAMHSI reported significant reductions in their child's symptoms in several areas: overall mental health symptoms (SDQ-P; d=0.67), emotional symptoms (SDQ-P; d=0.73), hyperactive symptoms (SDQ-P; d=0.46), impact score (SDQ-P; d=0.68), emotionality (EQ-P; d=0.57), and (TSCYC) symptoms of anger (d=0.65), arousal (d=0.66), and | Pernebo et al (2018) conducted a study in Sweden which compared psychotherapy (n=19) and psychoeducational interventions (n=31) for children and young people exposed to intimate partner violence. The mothers in the CBI reported a significant reduction in their child's emotional symptoms (SDQ-P; d=0.34), in total post-traumatic stress |
|  | **Quality rating:** High Quality |  | dissociation ($d=0.76$). Large effects were reported by the mothers in the CAMHSI for a decrease in depressive symptoms (TSCYC; $d=0.99$) and an increased capacity for emotion regulation (EQ-P; $d=0.85$)<br>**Additional Findings:**<br>The results of the study indicate that the psychotherapeutic intervention was somewhat more effective than the psych-educative intervention in reducing child symptoms in the aftermath of IPV. | (TSCYC; $d=0.35$), and in intrusive symptoms (TSCYC; $d=0.40$). Mothers in the CBI additionally reported a significant decrease in impact scores (SDQ-P; $d=0.62$). The mothers in the CAMHSI reported significant reductions in their child's symptoms in several areas: overall mental health symptoms (SDQ-P; $d=0.67$), emotional symptoms (SDQ-P; $d=0.73$), hyperactive symptoms (SDQ-P; $d=0.46$), impact score (SDQ-P; $d=0.68$), emotionality (EQ-P; $d=0.57$), and (TSCYC) symptoms of anger ($d=0.65$), arousal ($d=0.66$), and dissociation ($d=0.76$). Large effects were reported by the mothers in the CAMHSI for a decrease in depressive symptoms (TSCYC; $d=0.99$) and an increased capacity for emotion regulation (EQ-P; $d=0.85$) |
|  |  |  |  | (Pernebo et al., 2018). |
| **Pernebo et al (2019)**<br>(Pernebo et al., 2019)<br>**Sweden** | **Study Design:** The study used a quasi-experimental design with assessment before (T1) and after termination of (T2) the intervention.<br>**Type of intervention:** Psychotherapy and psychoeducational interventions<br>**Data collection methods:** Six measures were applied: (1) the revised Conflict Tactics Scales (CTS2; Straus, Hamby, Boney-McCoy, & Sugarman, 1996) to cover continuing levels of exposure to violence ($\alpha=0.59$ to $\alpha=0.90$); (2) the Strengths and Difficulties Questionnaire (SDQ-P; Goodman, Ford, Simmons, Gatward, & Meltzer, 2000) measuring child mental health ($\alpha=0.58$ to $\alpha=0.84$); (3) the Trauma Symptom Checklist for Young Children (TSCYC; Briere et al., 2001) to assess child symptoms of post-traumatic stress ($\alpha=0.74$ to $\alpha=0.91$); (4) the Emotion Questionnaire for parents (EQ-P; Rydell, Berlin, & Bohlin, 2003) for child emotionality and emotional regulation ($\alpha=0.85$); (5) the Brief Symptom Inventory (BSI; Derogatis & Melisaratos, | **Sample size:** n=31 in the psychoeducation group and n=19 CAMHS psychotherapy group<br>**Participants:** Children exposed to intimate partner violence<br>**Setting:** Community based intervention in Sweden<br>**Dates of data collection:** Unclear However, it was noted that it 'took 2 years to conduct this study'. However, this study included a baseline, 6-month, and 12-month follow-up). | **Primary Findings:** Mothers reported sustained, continuing, and additional significant improvements in children's symptoms of general psychological health and trauma symptoms from pre-intervention to the follow-up assessments. Significant improvements between post-assessment and 6-month follow-up in children's scores on the TSCYC on total post-traumatic stress ($p = 0.031$, $d = 0.33$), intrusion ($p = .033$, $d = 0.29$), avoidance ($p = .005$, $d = 0.64$), and dissociation ($p = .020$, $d = 0.64$) were reported by the mothers in the CAMHSI. No significant changes were reported between the 6- and 12-month follow-ups. Between the post-treatment assessment and the 12-month follow-up there were significant decreases in maternal report of child scores on the SDQ scale on emotional symptoms ($p = .004$, $d = 0.67$), the TSCYC scale on total post-traumatic stress ($p = 0.015$, $d = 0.44$), and avoidance ($p = 0.014$, $d=0.55$) for children in the CAMHSI. Between the post-treatment assessment and the 6-month follow-up there was a significant decrease in children's scores on the SDQ scale on anger ($p = .038$, $d = 0.42$) as reported by mothers in the CBI. Between the 6- and 12-month follow-ups there was a significant improvement in the CBI in maternal report of scores on the TSCYC scale on anxiety ($p = .023$, $d = 0.29$) and on the SDQ scale on prosocial behavior ($p = .044$, $d = 0.49$). Between the post-treatment assessment and the 12-month follow-up there were no significant | Pernebo et al (2019) (Pernebo et al., 2019) was a study including n=31 in the psychoeducation group and n=19 CAMHS psychotherapy group from community based interventions in Sweden.<br>Mothers reported sustained, continuing, and additional significant improvements in children's symptoms of general psychological health and trauma symptoms from pre-intervention to the follow-up assessments. Significant improvements between post-assessment and 6-month follow-up in children's scores on the TSCYC on total post-traumatic stress ($p = 0.031$, $d = 0.33$), intrusion ($p = .033$, $d = 0.29$), avoidance ($p = .005$, $d = 0.64$), and |
|  | 1983) to measure maternal mental health ($\alpha=0.93$); (6) the Impact of Event Scale—Revised (IES-R; Weiss, 2004) for maternal post-traumatic stress symptoms ($\alpha=0.79$ to $\alpha=0.90$).<br>**Quality rating:** High Quality |  | **Additional Findings:**<br>The results of the study indicate that the children did benefit from the two interventions studied and that the outcomes of reduced child symptoms and protection from exposure to violence were sustainable. Children with severe trauma symptoms benefited the most, though maternal psychological problems may for some have hindered recovery. | dissociation ($p = .020$, $d = 0.64$) were reported by the mothers in the CAMHSI. No significant changes were reported between the 6- and 12-month follow- ups. Between the post-treatment assessment and the 12-month follow-up there were significant decreases in maternal report of child scores on the SDQ scale on emotional symptoms ($p = .004$, $d = 0.67$), the TSCYC scale on total post-traumatic stress ($p = 0.015$, $d = 0.44$), and avoidance ($p = 0.014$, $d=0.55$) for children in the CAMHSI. Between the post-treatment assessment and the 6-month follow-up there was a significant decrease in children's scores on the SDQ scale on anger ($p = .038$, $d = 0.42$) as reported by mothers in the CBI. Between the 6- and 12-month follow-ups there was a significant improvement in the |
| | | | | CBI in maternal report of scores on the TSCYC scale on anxiety ($p = .023$, $d = 0.29$) and on the SDQ scale on prosocial behavior ($p = .044$, $d = 0.49$). Between the post-treatment assessment and the 12-month follow-up there were not significant. |
| **Schubert et al (2021)**<br>(Schubert, 2021)<br>**USA** | **Study Design:** Controlled trial<br>**Type of intervention:** 12-week group program delivered by domestic violence agency staff that provides psychoeducation on the impact of trauma and domestic violence and aims to improve parent and child well-being. The impact of the Child Witness to Domestic Violence (CWDV) program was tested in an intervention group ($n = 69$ children, 33 mothers) who participated in CWDV and control group ($n = 80$ children, 39 mothers) consisting of children whose mothers received adult-focused domestic violence services but were not enrolled in CWDV or other child-focused services.<br>**Data collection methods:** Strengths and Difficulties Questionnaire (SDQ) | **Sample size:** intervention group ($n = 33$ adults and 69 children aged 2 to 17), control group ($n = 39$ adults and 80 children aged 2 to 17)<br>**Participants:** Children and their mothers who were victims of domestic violence<br>**Setting:** Unclear. This study was not conducted with university or traditional research entity affiliated researchers.<br>**Dates of data collection:** the intervention (Spring 2017- Spring 2019) and the control (Winter 2016/2017- Summer 2018) | **Primary Findings:** Children who participated in CWDV demonstrated less hyperactivity, fewer negative emotional symptoms, and fewer total behavioural difficulties than their peers who did not participate in CWDV. Specifically, multiple regression analyses indicated that condition (intervention vs. control) was a significant predictor of child hyperactivity ($B = -.85$, $p = .06$; mean group difference at post-test = 0.63 out of 10), negative emotional symptoms ($B = -1.14$, $p = .01$; mean group difference at post-test = 1.22 out of 10), and total behavioural difficulties ($B = -2.48$, $p = .02$; mean group difference at post-test = 2.23 out of 40). Further,<br>**Additional Findings:** Mothers who participated in CWDV demonstrated higher hope pathways (i.e., their belief that they have the ability and means to achieve their goals) than mothers who received only adult focused DV services over the same amount of time. | Schubert (2021) conducted a trial in the USA which compared a child witness to domestic violence programme for mothers and children with a control group (Schubert, 2021). As a quasi-experimental design was used with different time periods for the intervention and the control, the possibility of selection effects was high. However, it was found that Children who participated in CWDV demonstrated less hyperactivity, fewer negative emotional symptoms, and fewer total behavioural difficulties than their peers who did not |
| | Dispositional Hope Scale (DHS)<br>**Quality rating:** Moderate Quality | | | participate in CWDV. Specifically, multiple regression analyses indicated that condition (intervention vs. control) was a significant predictor of child hyperactivity ($B = -.85$, $p = .06$; mean group difference at post-test = 0.63 out of 10), negative emotional symptoms ($B = -1.14$, $p = .01$; mean group difference at post-test = 1.22 out of 10), and total behavioural difficulties ($B = -2.48$, $p = .02$; mean group difference at post-test = 2.23 out of 40). |

**Abbreviations:** Kiddie Schedule for Affective Disorders and Schizophrenia, Present and Lifetime Version (K-SADS-PL); Child's behaviour. The Child Behaviour Checklist (CBCL); JPPSST = Joseph Pre-School and Primary Self-Concept Screening Test; The Revised Knox Preschool Play Scale (R-KPPS)

**Table 6.2.4.** Summary of clinical effectiveness of Play Therapy

| Citation (Country) | Study Details | Participants & setting | Key findings | Observations/notes |
| --- | --- | --- | --- | --- |
| **Waldman-Levi et al (2015)**<br>(Waldman-Levi and Weintraub, 2015) | **Study Design:** pre-test/post-test two-group control study design<br>**Type of intervention:** (n = 20 mother-child dyads) to the Family Intervention for Improving Occupational Performance (FI-OP) program and the control | **Sample size:** N=20 mother child dyads (children aged between 1 and 6 years old)<br>**Participants:** N=20 children (and mother) days in the intervention group and N=17 child and mother dyads in the control group | **Primary Findings:** After the intervention, mother-child interaction was significantly better in the FI-OP group than in the playroom group. The Mann-Whitney U test used to compare the study groups' difference scores (post-test vs. pre-test; see Table 4) with respect to children's playfulness (ToP) revealed no significant difference between | Waldman-Levi et al (2015) (Waldman-Levi and Weintraub, 2015) conducted a pre-test-post-test two-group control study design in Israel with twenty mother child dyads (children aged between 1 |
| **Israel** | group (n = 17 dyads) to a playroom program.<br>Both programs consisted of eight 30-minute sessions.<br>**Data collection methods:**<br>The Revised Knox Preschool Play Scale (R-KPPS) and The Test of Playfulness (ToP) (observational tool)<br>Parent child interactions were also observed (Coding Interactive Behaviour (CIB) rating system.<br>**Quality rating:** Moderate Quality | **Setting:** Community settings in Israel (shelters)<br>**Dates of data collection:** Unclear (not provided in the publication) | the FI–OP and playroom groups. The Wilcoxon test examining within-group differences in child’s playfulness showed no significant difference in the FI–OP group, $Z = -1.19$, $p > .05$, but a significant difference in the playroom group, $Z = -1.76$, $p < .05$. The results of this study indicate that children in the FI–OP program significantly improved their play skills compared with the playroom group, in which no significant improvement was noted.<br>**Additional Findings:**<br>The creation of a safe space during the intervention may facilitate mother-child interaction, however, there was no follow up phase in this study, limiting the impact of the positive results for the eight sessions. | and 6 years old). The intervention aimed to improve occupational performance and there was also a playroom program which acted as the control arm. After the intervention, mother–child interaction was significantly better in the FI–OP group than in the playroom group. The Mann–Whitney U test used to compare the study groups’ difference scores (post-test vs. pre-test; see Table 4) with respect to children’s playfulness (ToP) revealed no significant difference between the FI–OP and playroom groups. The Wilcoxon test examining within-group differences in child’s playfulness showed no significant difference in the FI–OP group, $Z = -1.19$, $p > .05$, but a significant difference in the playroom group, $Z = -1.76$, $p < .05$. The results of this study indicate that children in the FI–OP program significantly improved their play skills compared with the playroom group, in which no significant improvement was noted. |

**Table 6.2.5.** Summary of clinical effectiveness of Parenting Skills Training

| **Citation (Country)** | **Study Details** | **Participants & setting** | **Key findings** | **Observations/notes** |
| --- | --- | --- | --- | --- |
| **Jouriles et al (2001)**<br>(Jouriles et al., 2001)<br>**USA** | **Study Design:** Randomised controlled study<br>**Type of intervention:** Child management skills to mothers (Psycho-educational intervention). Control: usual care<br>**Data collection methods:** Child Behaviour Checklist (CBCL)<br>**Quality rating:** Low Quality | **Sample size:** 36 families (mothers and children with at least one child with a conduct disorder (oppositional defiance disorder) between 4 and 9 years old).<br>**Participants:** Children 4-9 years old. 26 boys and 10 girls.<br>**Setting:** Community settings in the USA<br>**Dates of data collection:** Unclear (not reported in the paper) | **Primary Findings:**<br>The results evaluating the children over time showed a significant interaction ($p < .01$) on the CBCL checklist. The children in the treatment condition improved at a faster rate (slope -3.53) than the children in the control condition (slope = -.95). Children were assessed 5 times in all, and by assessment 3, there was no difference between the groups).<br>**Additional Findings:**<br>The intervention's aim was to support mothers during the transition from a shelter for women who have experienced domestic violence, teaching women to effectively address their' children' conduct problems during this transitional period. The outcomes for mothers were also positive. | Jouriles et al (2001) investigated a psycho-educational intervention and found that although the behaviour of the children in both intervention and control groups improved, the intervention children's behaviour improved at a faster rate (slope - 3.53) than the children in the control condition (slope = -.95). Children were assessed 5 times in all, and by assessment 3, there was no difference between the groups. |
| **Jouriles et al (2009)**<br>(Jouriles et al., 2009)<br>**USA** | **Study Design:** Randomised controlled study<br>**Type of intervention:** Child management skills to mothers (Psycho-educational intervention). Control: usual care<br>**Data collection methods:** Child Behaviour Checklist (CBCL) and Eyberg Child Behaviour Inventory for Observational oppositional behaviour. | **Sample size:** 66 families (with children between 4 and 9 years old) were recruited from shelters for women victims of domestic abuse.<br>**Participants:** 66 families including mothers and children (n=32 project support and n=34 comparison group)<br>**Setting:** Community settings in the USA<br>**Dates of data collection:** October 1996 to January 2000. | **Primary Findings:**<br>During the intervention period, child conduct problems decreased in the Project Support group ($p < .001$) as well as in the comparison group ($P < .01$). However, they decreased more rapidly in the Project Support group than in the comparison group ($P < .01$). For the follow-up period, conduct problems continued to decrease in the Project support group ($p < .005$) but not in the comparison group (NS).<br>**Additional Findings:** | **Jouriles et al (2009)**<br>(Jouriles et al., 2009) returned to their original 2001 published work with mothers and children recruited from shelters, with a bigger sample and found similar results. During the intervention period, child conduct problems decreased in the Project Support group |
|  | **Quality rating:** High Quality |  | The effectiveness of the Project Support intervention is dependent on the mother's general well-being and parenting skills. | (P <.001) as well as in the comparison group (P <.01). However, they decreased more rapidly in the Project Support group than in the comparison group (P <.01). For the follow-up period, conduct problems continued to decrease in the Project support group (P < .005) but not in the comparison group (NS). The effectiveness of the Project Support intervention is dependent on the mother's general well-being and parenting skills. |

**Table 6.2.6.** Summary of cost-effectiveness/cost analysis studies of interventions to improve outcomes for children and young people who had witnessed domestic violence

| **Citation (Country)** | **Study Details** | **Participants & setting** | **Key findings** | **Observations/notes** |
| --- | --- | --- | --- | --- |
| **ADVA. Community Perpetrator Programme REPAIR (2009)**<br>**England** | **Study Design:** Evaluation<br>**Type of intervention:** Advocacy and psychoeducation<br>**Data collection methods:** Focus groups and questionnaires<br>**Quality rating:** Quality Unclear* | **Sample size:** Fathers, mother, and children on the ADVA Community Perpetrator Programme REPAIR<br>**Participants:** 20 children of fathers on the perpetrator programme (aged between 5 and 18 years).<br>**Setting:** Community setting in England | **Primary Findings:** The evaluation of REPAIR measured uptake, retention and costs specifically looking at mental health, behaviour, and family outcomes. The majority of the 20 children of fathers on the perpetrator programme demonstrated decreased anxiety, stress and anger and an improved relationship with mothers and peers as reported by the mothers. | The REPAIR perpetrator programme (Sue Penna Associates, 2009) which was established in three areas of Devon focused on motivation, responsibility, safety, and acknowledgement |
| (Sue Penna Associates, 2009) | *Unclear due to inappropriate appraisal tools. | **Dates of data collection:**<br>2005-2008 | **Additional Findings:**<br>The overall annual cost of the REPAIR programme was £186,390. The REPAIR programme would serve 24 families per year (including mother and father and children) (Sue Penna Associates, 2009). | for men. The women's services were based on individual needs where a woman's support worker provided advocacy, practical and psychological support, and the children's groups focused on safety, risk assessment, the development of resilience, appropriate coping strategies and support networks and processing difficult feelings, along with the element of liaising closely with the school and especially with the classroom teacher.<br>The majority of the 20 children of fathers on the perpetrator programme demonstrated decreased anxiety, stress and anger and an improved relationship with mothers and peers as reported by the mothers.<br>The overall annual cost of the REPAIR programme was £186,390. The REPAIR programme would serve 24 families per year<br>(including mother and father and children) (Sue Penna Associates, 2009). |
| **Howarth et al (2016)** | **Study Design:** Cost-effectiveness<br>**Type of intervention:** Various cognitive and behavioural interventions<br>**Data collection methods:** RCTs<br>**Quality rating:** | **Sample:** 8 studies with mostly small samples (fewer than n=80 participants in most studies).<br>**Participants:** Children and young people and their parents who have been exposed to domestic violence and abuse<br>**Setting:** USA<br>**Dates of data collection:** Up to 2015 | **Primary Findings:**<br>For behavioural outcomes, a psychoeducational intervention delivered to parent and child in parallel [type C: PEd(C + P)] is likely to be cost-effective among the interventions that we compared (ICER = 3722 per standardized mean difference). | Cost-effectiveness analysis by Howarth et al (2016) suggested that for behavioural outcomes, a psychoeducational intervention delivered to parent and child in parallel is likely to be cost-effective among the interventions that they compared if willingness to pay was approximately £8000 (ICER = 3722 per Standard Mean Difference (SMD)).<br>For mental health outcomes, it is very likely that a psychoeducational intervention delivered to the child would be cost-effective. If willingness to pay per SMD in mental health outcomes is high (ICER > £22,575/SMD), cognitive-behavioural therapy (delivered to the parent, child and dyad) may be equally cost-effective. |
|  |  |  |  | Costs were estimated based on the reported description of the interventions in the study publications. They found much heterogeneity in the data that reflected the complex nature of these interventions. Training costs were not included in the analysis before this would be a one-off cost and not an annual cost for rolling interventions. The variety of intervention venues were not costed either and neither were ongoing supervision costs. Therefore, there was a large degree in uncertainty about the intervention costs, which is a limitation of this cost-effectiveness analysis.<br>Howarth et al (2016) noted that their analyses were intended to be 'hypothesis-generating' to inform the future design of research studies, rather than robust estimates of effectiveness and cost-effectiveness. |
|  |  |  |  | Therefore, their tentative conclusions concerning which interventions to pursue in future research studies should be treated with caution. |
| **Nolas, (2012)**<br>(Nolas et al., 2012)<br>**England** | **Study Design:** Evaluation including cost of group therapy.<br>**Type of intervention:** Group therapy intervention<br>**Data collection methods:** Qualitative interviews and cost analysis of group therapy sessions.<br>**Quality rating:** Quality Unclear*<br>*Unclear due to inappropriate appraisal tools. | **Sample size:** 24 children between 4 and 21 years.<br>**Participants:** 24 children were involved in the study<br>**Setting:** Community setting in England. The group therapy was a 12-week group intervention for children and young people (aged 4–21 years) in recovery from domestic violence and abuse (DVA), alongside a concurrent group work programme for their mothers.<br>**Dates of data collection:** Unclear, but pre-publication of the evaluation report in 2012, but otherwise not noted in the methods section. | **Primary Findings:**<br>The estimated cost of running a 12- week group was £9,123.96 (per group), which equated to £1,303.25 per child (with a maximum of 7 children in the group).<br>**Additional Findings:**<br>The costs included training costs, one-off costs, essential costs such as room hire and staff costs and additional costs such as transport, interpreter, and evaluation costs. | In the Nolas (2012) evaluation including costing, it was found that the costs of having 7 children in a community group therapy session for 12 weeks was a little over £1,300 in 2012 (this would be £1,725 in December 2022 (Bank of England Inflation Calculator). Sex balance in the children's groups was important to children and they valued attending separate groups from their siblings. For a minority of children, the timings of the groups had been inconvenient, as they missed out on school curricular and extracurricular activities (Nolas et al., 2012). |
| **Sharp et al (2011)**<br>(Sharp et al., 2011) | **Study Design:** Cost analysis.<br>**Type of intervention:** 12-week group therapy (CEDAR Project) (an early intervention with a | **Sample size:** A total of 27 children and young people, aged five to 16 years were interviewed, both genders were also well represented. | **Primary Findings:**<br>**Intervention:**<br>Children's accounts of being involved in groupwork were positive on the whole. Children's ability to participate effectively in | Sharp et al (2011) conducted a study in Scotland UK and found that the overall costs of the pilot was |
|  | combination of play and discussion). Topics covered were: behaviour, knowledge of safety, attitudes to violence, relationships (making connections) and mental health.<br>**Data collection methods:**<br>Qualitative interviews<br>**Quality rating:** Quality Unclear*<br>*Unclear due to inappropriate appraisal tools. | **Participants:** Children and young people<br>**Setting:** Community settings in Scotland<br>**Dates of data collection:** January 2009-March 2011 | groupwork varied according to individual inclination, the degree of behavioural issues and learning difficulties/physical disability. Fear of being identified as having suffered from domestic abuse can be a deterrent factor. In terms of outcomes, groupwork programmes have shown positive changes in children's aggressive behaviour; greater knowledge of safety; changed attitudes to violence; strengthened relationships between mothers and children; lowering of anxiety and increased self-esteem. Groupwork also facilitated access to trusted networks of informal support and friends.<br>**Cost implication:**<br>The overall costs of the pilot were approximately £0.83m over the three-year period. There were 39 groups held during this time.<br>**Additional Findings:**<br>Children expressed that they liked attending the group sessions. | approximately £0.83m over the three-year period (Sharp et al., 2011).<br>A full assessment of value for money would need to consider all the short and longer-term costs, the direct and indirect costs and the potential for savings, even if they may not be directly or immediately quantifiable. Data available was limited to short-term service delivery direct costs. Cedar coordinators' salary costs were the largest single item. |
| **Aas (2019)**<br>(Aas et al., 2019)<br>**Finland** | **Study Design:** RCT and cost-effectiveness<br>**Type of intervention:**<br>Trauma Focused Cognitive Behavioral Therapy (TF-CBT)<br>Control: Treatment as usual (TAU)<br>**Data collection methods:**<br>15D questionnaire<br>**Quality rating:** High Quality | **Sample size:** 123 youths from 10 to 18 years<br>**Participants:**<br>the majority were girls (n = 124, 79.5%) with a mean age of 15.1 years.<br>**Setting:** community clinics (Finnish population).<br>**Dates of data collection:** between April 2008 and July 2013 | **Primary Findings:**<br>ICER was 140, indicating that TF-CBT both reduced the resource use and increased health gains<br>TF-CBT is likely to be cost-effective and hence should be implemented as guideline treatment for adolescents with post-traumatic stress disorder<br>**Additional Findings:** | Aas (2019) conducted a RCT and cost-effectiveness study to investigate the effectiveness and cost-effectiveness of a CBT intervention focused on trauma in Finland (Aas et al., 2019). It was found that for every Quality-of-Life Year (QALY) gained, about |
|  |  |  | HRQoL increased for both treatment groups and there are no significant differences in QALYs.<br>Resource use is significantly higher in the TF-CBT group for minutes per session, while total minutes of therapy are significantly lower in this group.<br>Psychological counseling services, welfare services and medication, 3 are lower in the TF-CBT group post-treatment<br>Cost-Effectiveness Analysis Curve (CEAC): 87% | *126 hours of therapy was saved.*<br>*The use of other services, such as welfare services and school nurse, were also lower for the TF-CBT group compared to the treatment as usual group.*<br>*Based on the sensitivity analysis, depending on the threshold for a QALY gained, the likelihood of TF-CBT being cost-effective varied between 0.87 and 0.95.* |
| **Shearer (2018)**<br>(Shearer et al., 2018)<br>**England** | **Study Design:** RCT and cost-effectiveness<br>**Type of intervention:** cognitive therapy<br>**Data collection methods:** Strengths and Difficulties Questionnaire (SDQ)<br>**Quality rating:** Moderate Quality | **Sample size:** There were n=14 young people in the treatment group, and n=15 in the control group.<br>**Participants:** Children and adolescents were included if they were aged 8–17 years<br>**Setting:** Emergency Departments, community mental health teams, primary care, schools and other health clinics across the East of England, UK.<br>**Dates of data collection:** Unclear 11-week randomized waiting list controlled trial | **Primary Findings:**<br>The 3-year ICER was £2,250 per QALY which is well below the NICE threshold of between £20,000 and £30,000 per QALY<br>**Additional Findings:**<br>The treatment effect was significant and treated patients gained more QALYs than untreated ones (2.370, 2.324). Difference (0.0577).<br>The CEAC was between 60% and 69%. | Shearer (2018) investigated the effectiveness and cost effectiveness of a CBT intervention in the East of England (Shearer et al., 2018). This study provided preliminary evidence for the cost-effectiveness of cognitive therapy for children and young people who had the intervention was also cost effective at £2,250 per QALY. |

**Abbreviations:** Randomised Controlled Trial (RCT); Child and Adolescent Mental Health Service (CAMHS); Child Witness to Domestic Violence (CWDV) program

### 6.3 Quality appraisal tables

Tables 6.3.1-6.3.3 are critical appraisal tables for the included studies in this rapid review.

**Table 6.3.1.**
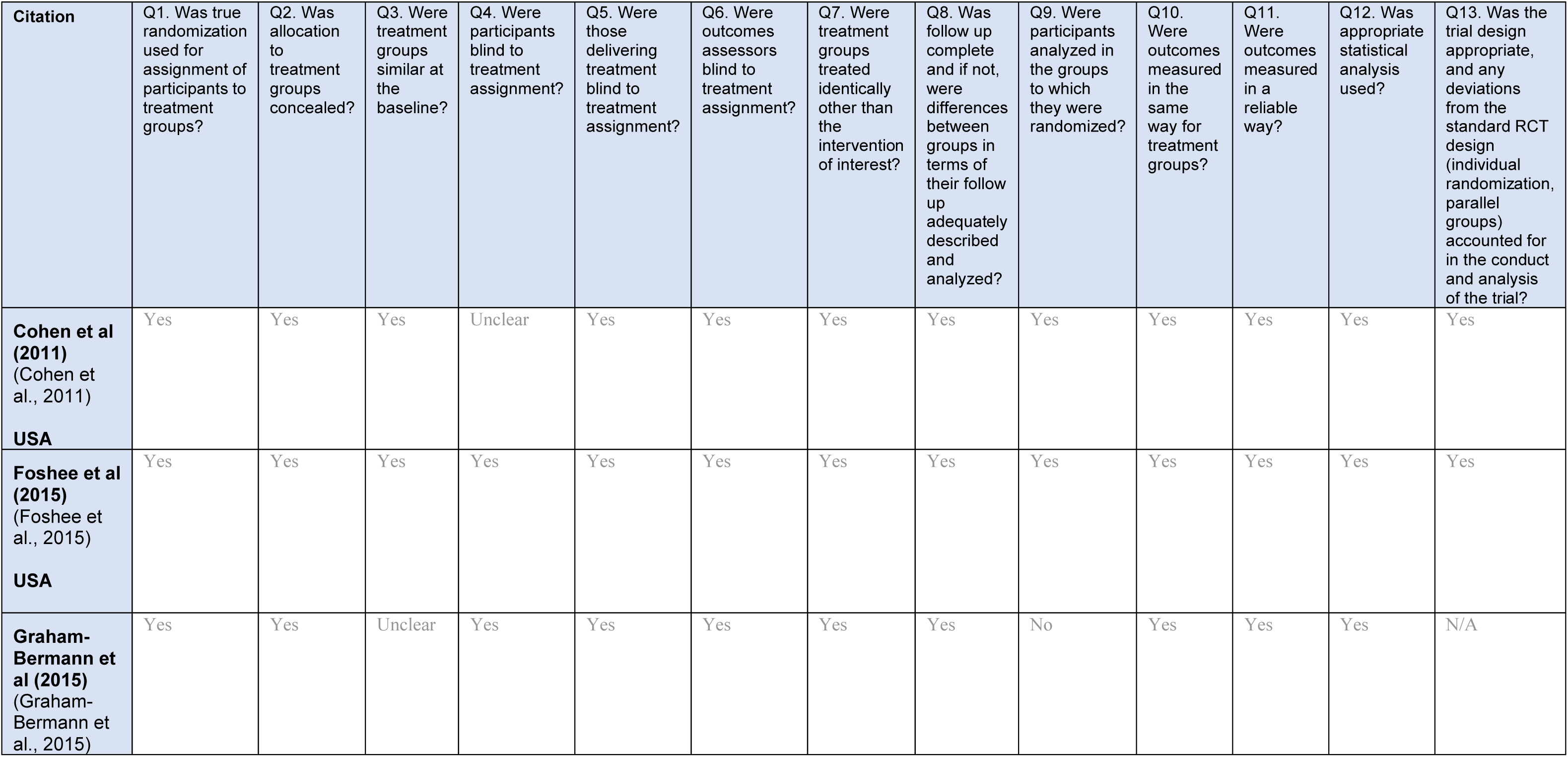
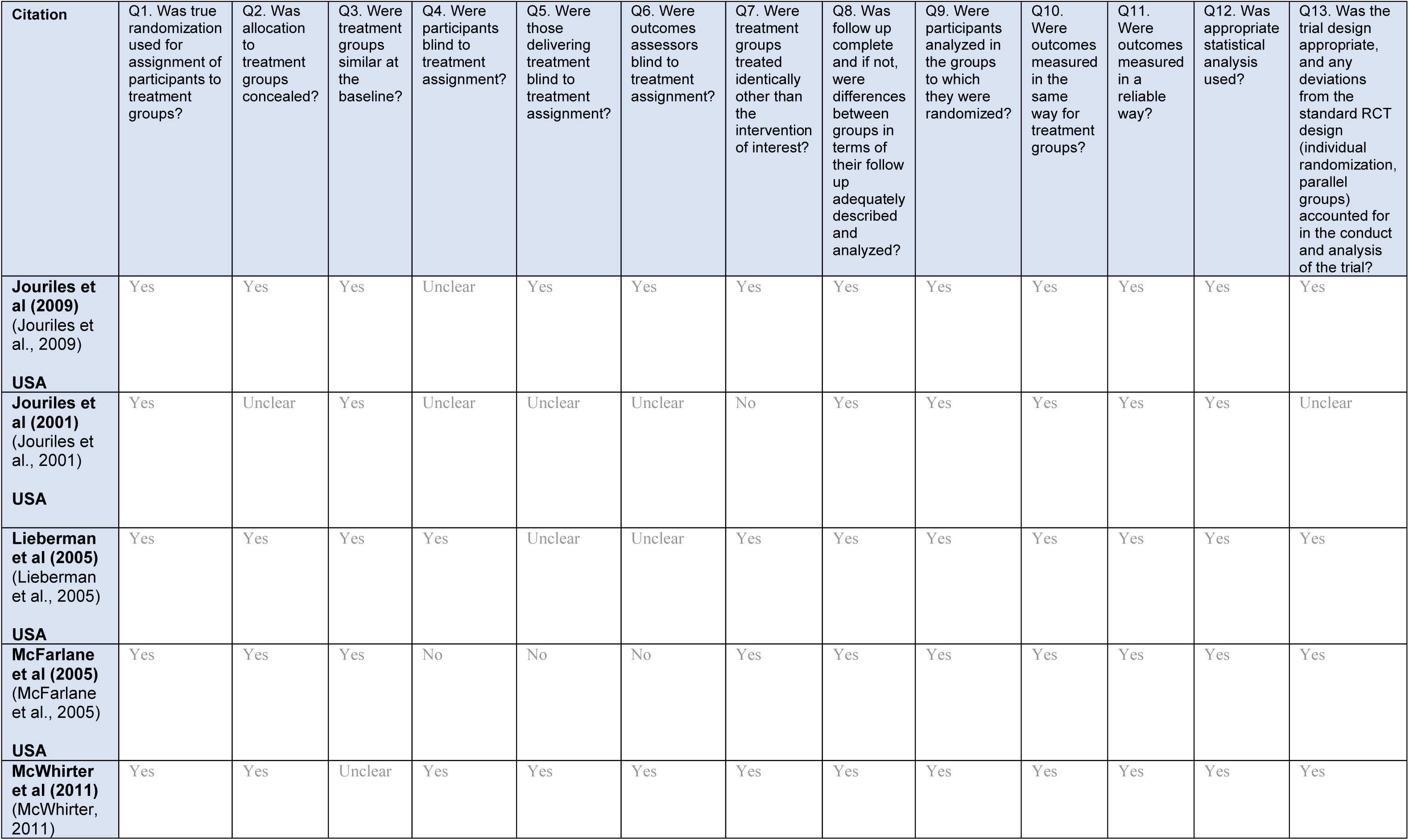
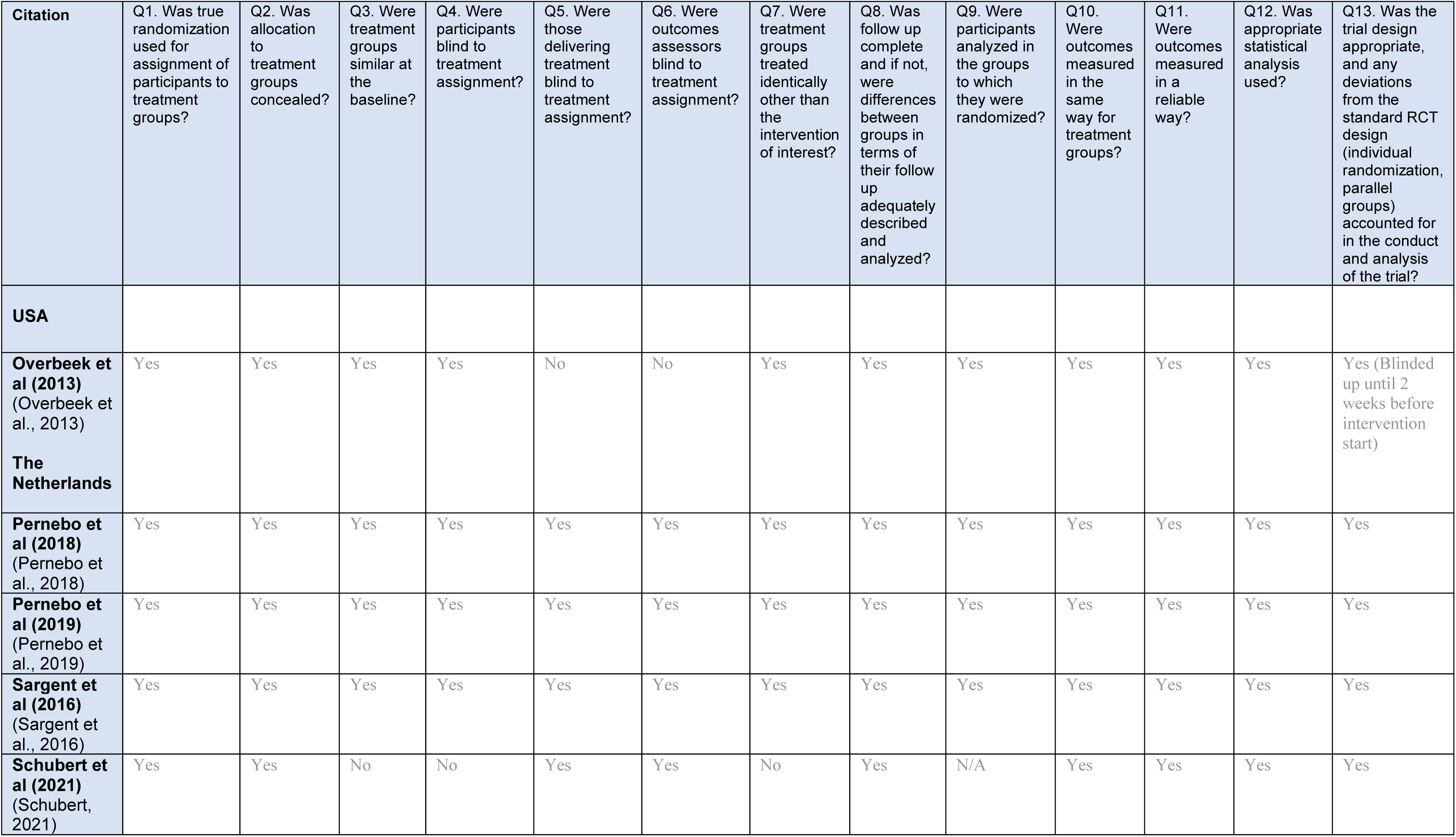
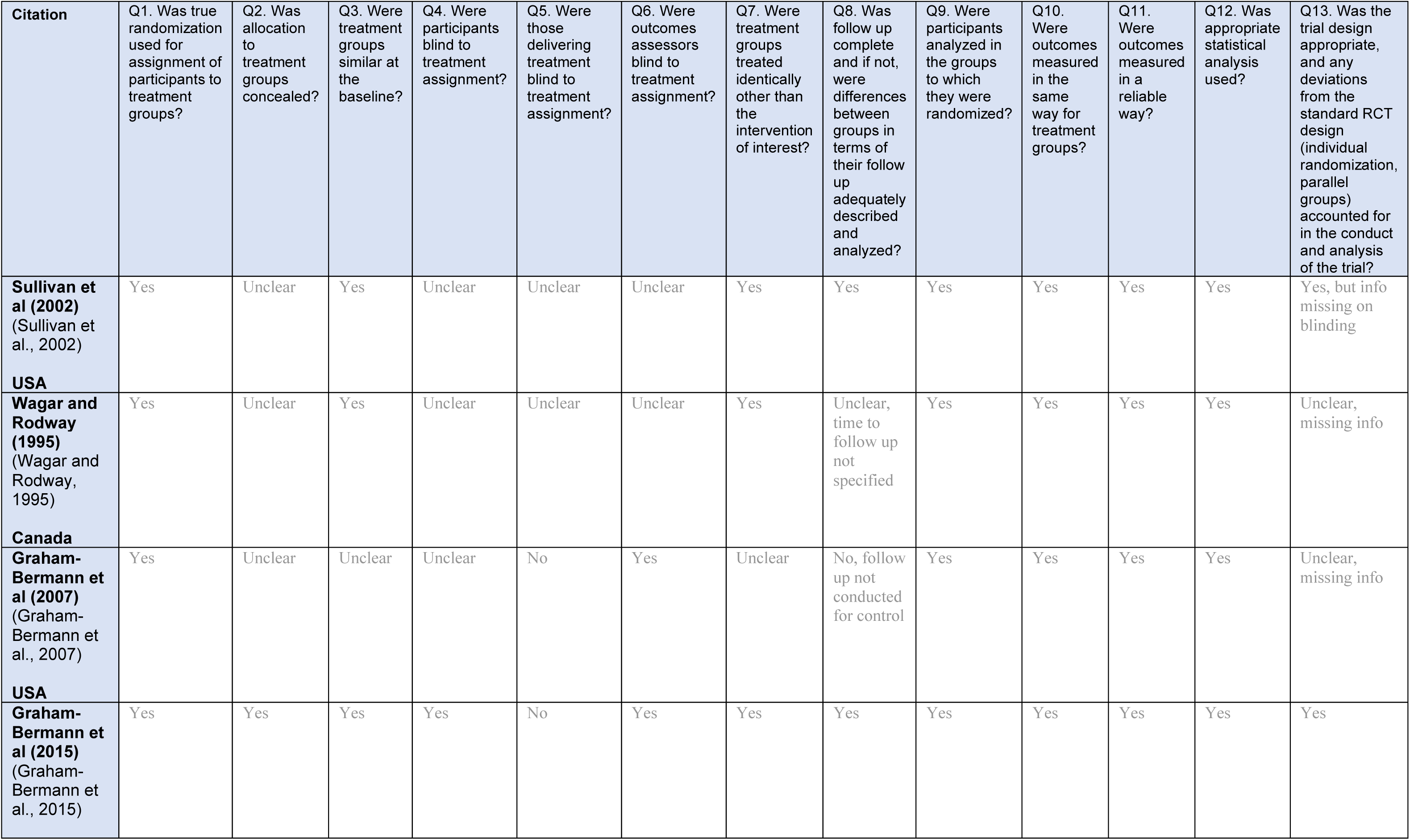
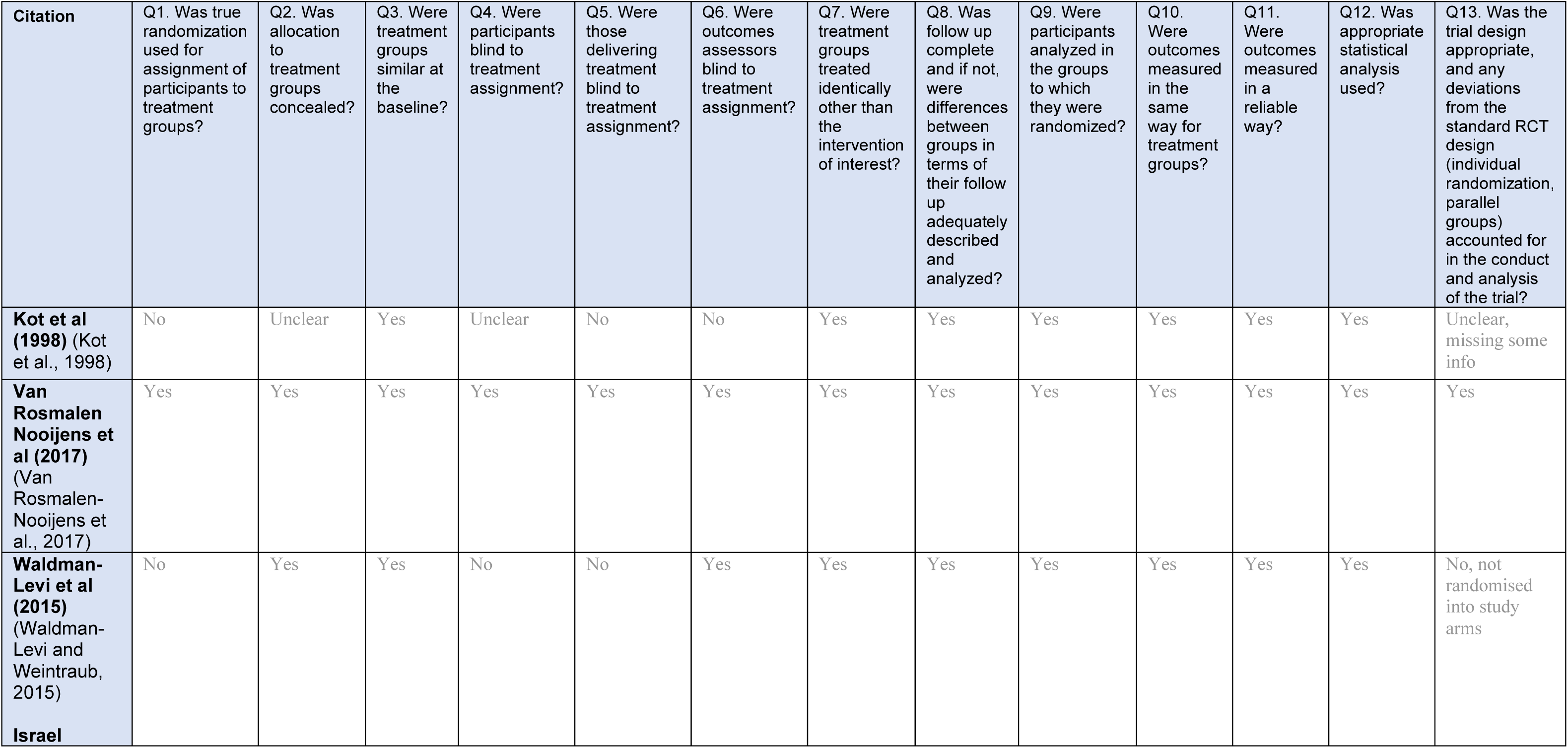
JBI Critical Appraisal Checklist for Randomised Control Trials

**Table 6.3.2.** CHEERS reporting guidance checklist for Health Economic Evaluations (Husereau et al., 2022).

| Citation* | 1 | 2 | 3 | 4 | 5 | 6 | 7 | 8 | 9 | 10 | 11a | 11b | 12 | 13a | 13b | 14 | 15 | 16 | 17 | 18 | 19 | 20a | 20b | 21 | 22 | 23 | 24 | Score |
| --- | --- | --- | --- | --- | --- | --- | --- | --- | --- | --- | --- | --- | --- | --- | --- | --- | --- | --- | --- | --- | --- | --- | --- | --- | --- | --- | --- | --- |
| Aas et al (2019)<br>(Aas et al., 2019) | Y | Y | Y | Y | Y | Y | Y | Y | Y | Y | Y | Y | Y | Y | Y | Y | Y | Y | Y | Y | Y | Y | Y | Y | Y | Y | Y | High |
| Adva (2009)<br>(Sue Penna Associates , 2009) | N | Y | Y | N | Y | Y | N | N | Y | N | N/A | N/A | N/A | N | N | Y | N/A | N/A | N | N/A | Y | N/A | N/A | N/A | Y | Y | N | Unclear due to inappropriate appraisal tools |
| Nolas et al<br>(Nolas et al., 2012) | N/A | Y | Y | N | Y | N | Y | N | N | N | N/A | Y | N/A | N/A | N/A | Y | N/A | N/A | Y | N/A | N | N/A | N/A | N/A | Y | Y | N | Unclear due to inappropriate appraisal tools |
| Sharp et al (2011)<br>(Sharp et al., 2011) | N/A | Y | Y | Y | Y | N | Y | N | N | N | N/A | N/A | N/A | N/A | N/A | N/A | N/A | N/A | N | N/A | N/A | N/A | N/A | N/A | Y | N | N | Unclear due to inappropriate appraisal tools |
| Shearer et al (2018)<br>(Shearer et al., 2018) | Y | Y | Y | N | Y | Y | Y | Y | N | Y | Y | Y | Y | Y | Y | Y | Y | Y | Y | Y | Y | Y | Y | Y | Y | Y | Y | Moderate |

\*CHEERS Checklist questions are shown in the table below.

| Section/item | Item No | Recommendation | Reported on page No/ line No |
| --- | --- | --- | --- |
| **Title and abstract** |  |  |  |
| **Title** | 1 | Identify the study as an economic evaluation or use more specific terms such as “cost-effectiveness analysis”, and describe the interventions compared. |  |
| **Abstract** | 2 | Provide a structured summary of objectives, perspective, setting, methods (including study design and inputs), results (including base case and uncertainty analyses), and conclusions. |  |
| **Introduction** |  |  |  |
| **Background and objectives** | 3 | Provide an explicit statement of the broader context for the study. Present the study question and its relevance for health policy or practice decisions. |  |
| **Methods** |  |  |  |
| **Target population and subgroups** | 4 | Describe characteristics of the base case population and subgroups analysed, including why they were chosen. |  |
| **Setting and location** | 5 | State relevant aspects of the system(s) in which the decision(s) need(s) to be made. |  |
| **Study perspective** | 6 | Describe the perspective of the study and relate this to the costs being evaluated. |  |
| **Comparators** | 7 | Describe the interventions or strategies being compared and state why they were chosen. |  |
| **Time horizon** | 8 | State the time horizon(s) over which costs and consequences are being evaluated and say why appropriate. |  |
| **Discount rate** | 9 | Report the choice of discount rate(s) used for costs and outcomes and say why appropriate. |  |
| **Choice of health outcomes** | 10 | Describe what outcomes were used as the measure(s) of benefit in the evaluation and their relevance for the type of analysis performed. |  |
| **Measurement of effectiveness** | 11a | Single study-based estimates: Describe fully the design features of the single effectiveness study and why the single study was a sufficient source of clinical effectiveness data. |  |
|  | 11b | Synthesis-based estimates: Describe fully the methods used for identification of included studies and synthesis of clinical effectiveness data. |  |
| **Measurement and valuation of preference-based outcomes** | 12 | If applicable, describe the population and methods used to elicit preferences for outcomes. |  |
| **Estimating resources and costs** | 13a | Single study-based economic evaluation: Describe approaches used to estimate resource use associated with the alternative interventions. Describe primary or secondary research methods for valuing each resource item in terms of its unit cost. Describe any adjustments made to approximate to opportunity costs. |  |
|  | 13b | Model-based economic evaluation: Describe approaches and data sources used to estimate resource use associated with model health states. Describe primary or secondary research methods for valuing each resource item in terms of its unit cost. Describe any adjustments made to approximate to opportunity costs. |  |
| **Currency, price date, and conversion** | 14 | Report the dates of the estimated resource quantities and unit costs. Describe methods for adjusting estimated unit costs to the year of reported costs if necessary. Describe methods for converting costs into a common currency base and the exchange rate. |  |
| **Choice of model** | 15 | Describe and give reasons for the specific type of decision analytical model used. Providing a figure to show model structure is strongly recommended |  |
| **Assumptions** | 16 | Describe all structural or other assumptions underpinning the decision-analytical model. |  |
| **Analytical methods** | 17 | Describe all analytical methods supporting the evaluation. This could include methods for dealing with skewed, missing, or censored data; extrapolation methods; methods for pooling data; approaches to validate or adjust (such as half cycle corrections) to a model; and methods for handling population heterogeneity and uncertainty. |  |
| **Results** |  |  |  |
| **Study parameters** | 18 | Report the values, ranges, references, and, if used, probability distributions for all parameters. Report reasons or sources for distributions used to represent uncertainty where appropriate. Providing a table to show the input values is strongly recommended. |  |
| **Incremental costs and outcomes** | 19 | For each intervention, report mean values for the main categories of estimated costs and outcomes of interest, as well as mean differences between the comparator groups. If applicable, report incremental cost-effectiveness ratios. |  |
| **Characterising uncertainty** | 20a | of sampling uncertainty for the estimated incremental cost and incremental effectiveness parameters, together with the impact of methodological assumptions (such as discount rate, study perspective). |  |
|  | 20b | Model-based economic evaluation: Describe the effects on the results of uncertainty for all input parameters, and uncertainty related to the structure of the model and assumptions. |  |
| **Characterising heterogeneity** | 21 | If applicable, report differences in costs, outcomes, or cost-effectiveness that can be explained by variations between subgroups of patients with different baseline characteristics or other observed variability in effects that are not reducible by more information. |  |
| **Discussion** |  |  |  |
| **Study findings, limitations, generalisability, and current knowledge** | 22 | Summarise key study findings and describe how they support the conclusions reached. Discuss limitations and the generalisability of the findings and how the findings fit with current knowledge. |  |
| **Other** |  |  |  |
| **Source of funding** | 23 | Describe how the study was funded and the role of the funder in the identification, design, conduct, and reporting of the analysis. Describe other non-monetary sources of support. |  |
| **Conflicts of interest** | 24 | Describe any potential for conflict of interest of study contributors in accordance with journal policy. In the absence of a journal policy, we recommend authors comply with International Committee of Medical Journal Editors recommendations. |  |

### 6.4 Information available on request

The study protocol is available on request from the first author of this rapid review report.

## 7.2 Acknowledgements

We express a huge debt of gratitude to the stakeholders, Mr Paul Webb, Dr Sally Anstey and Ms Alison Plant for their time, expertise and valuable input in guiding the outline of this rapid review report and for their assistance in identifying relevant outcomes.

## 8. ABOUT THE HEALTH AND CARE RESEARCH WALES EVIDENCE CENTRE

The Health and Care Research Wales Evidence Centre integrates with worldwide efforts to synthesise and mobilise knowledge from research.

We operate with a core team as part of [Health and Care Research Wales](https://healthandcareresearchwales.org/about-research-community/wales-covid-19-evidence-centre), Welsh Government, and are led by [Professor Adrian Edwards of Cardiff University](https://www.cardiff.ac.uk/people/view/123022-edwards-adrian).

The core team of the centre works closely with collaborating partners in the [Bangor Institute for Health and Medical Research (BIHMR)](https://www.bangor.ac.uk/medical-health-sciences/research), Bangor University, which includes the [Centre for Health Economics and Medicines Evaluation (CHEME)](https://cheme.bangor.ac.uk/) working in collaboration with Health and Care Economics Cymru, [Health Technology Wales](https://healthtechnology.wales/), [Public Health Wales Evidence Service](https://phw.nhs.wales/services-and-teams/observatory/evidence/), [Population Data Science, Swansea University](https://popdatasci.swan.ac.uk/about-us/) using [SAIL Databank](https://saildatabank.com/), the [Wales Centre for Evidence Based Care (WCEBC)](https://www.cardiff.ac.uk/healthcare-sciences/about-us/engagement/the-wales-center-for-evidence-based-care-wcebc), the [Specialist Unit for Review Evidence (SURE)](https://www.cardiff.ac.uk/specialist-unit-for-review-evidence) and [CASCADE](https://cascadewales.org/), Cardiff University.

**Director:**

Professor Adrian Edwards

**Website:** www.researchwalesevidencecentre.co.uk

## APPENDIX

## APPENDIX 1 Resources searched during Rapid Review Searching

The evidence presented in this rapid review were from the sources indicated in Table A.1.

**Table A.1:** Resources searched

| Resource | Number of hits |
| --- | --- |
| Howarth et al (2016) Systematic Review (Howarth et al., 2016) | 16 studies (between 1995 and 2015). |
| Howarth et al (2016) Cost-effectiveness (Howarth et al, 2016) | 1 |
| Health and Social Science Databases | (Excluding duplicates) |
| • Medline<br>• ASSIA<br>• PsycINFO<br>• CINAHL<br>• Embase<br>• Cochrane Library<br>• NIHR Centre for Reviews and Dissemination (CRD) database including NHS EED database | 7 (primary studies since 2015) |
| Additional resources searched |  |
| Google Advanced Search | 2 (cost-effectiveness studies) |
| Google | 3 (guidelines) |
| **Total** | **29** |

